# Hepatic safety of pretomanid- and pyrazinamide-containing regimens in TB Alliance clinical trials

**DOI:** 10.1101/2025.03.20.25324141

**Authors:** Jerry Nedelman, Mengchun Li, Morounfolu Olugbosi, Rebecca Bruning-Barry, Jeffrey Ambroso, Muge Cevik, Stephen Gillespie, Derek J Sloan, Maria Beumont, Eugene Sun

## Abstract

**Background:** In STAND and SimpliciTB, clinical trials for drug-susceptible tuberculosis (TB), regimens containing pretomanid, pyrazinamide, and other agents (PaZX) had more hepatotoxicity than the standard-of-care regimen of isoniazid, rifampicin, pyrazinamide, and ethambutol (HRZE). In Nix-TB and ZeNix, clinical trials for drug-resistant TB, the regimen of bedaquiline, pretomanid, and linezolid (BPaL) demonstrated a favorable benefit-risk profile. We compare the hepatic safety of HRZE, PaZX, and BPaL in their respective populations.

**Methods:** In this post-hoc analysis of data from six clinical trials, rates of treatment-emergent elevations of alanine transaminase (ALT) during the first eight weeks of treatment were estimated by Kaplan-Meier (KM) analysis and compared via log-rank testing and Cox modeling.

**Results:** The KM-estimated probabilities of treatment-emergent ALT elevations greater than 3 times the upper limit of normal (>3xULN) were 5.36%, 12.7%, and 11.4% for HRZE, PaZX, and BPaL, respectively, with the only significant (p < 0.05) difference being HRZE versus PaZX. The probabilities of ALT elevations >8xULN were 2.68%, 4.58%, and 1.05%, with the only significant difference being PaZX versus BPaL.

**Conclusions:** BPaL and HRZE have similar hepatic safety profiles in their respective populations. Pretomanid and pyrazinamide should be co-administered only when the benefit outweighs the risk.

## INTRODUCTION

Pretomanid is a nitroimidazole developed by TB Alliance. In 2019, the United States Food and Drug Administration approved pretomanid (Pa) at 200 mg once daily as part of a three-drug, six-month regimen with bedaquiline (B) and linezolid (L) to treat adults with pulmonary extensively drug-resistant tuberculosis (XDR-TB, pre-2021 definition) or intolerant or nonresponsive multidrug-resistant TB (MDR-TB)^1^. The regimen is referred to as BPaL. In December 2022, the World Health Organization recommended the use of a six-month treatment regimen composed of BPaL with or without moxifloxacin (M) rather than the nine-month or longer regimens in patients with MDR-TB and rifampicin-resistant TB^2^.

Promising results from phase 2b studies^3,4^ of pretomanid combinations prompted exploration of treatment shortening. The STAND^5^ trial tested pretomanid with moxifloxacin and pyrazinamide (Z) (PaMZ) versus the standard of care (isoniazid, rifampin, pyrazinamide, and ethambutol (HRZE)) in drug-susceptible TB (DS-TB). STAND was paused following three deaths in the PaMZ arms associated with hepatotoxicity. After review of the safety data, the Safety Monitoring Committee recommended resuming enrollment, but TB Alliance decided instead to pursue what appeared to be a more promising regimen, BPaMZ, in the SimpliciTB^6^ trial. BPaMZ demonstrated more rapid sputum culture conversion but a higher rate of withdrawals due to elevated hepatic enzymes.

After approval of Pa in the BPaL regimen, which was based on the Nix-TB^7^ study, TB Alliance conducted the ZeNix study^8^, which further optimized the linezolid dose. In both studies with BPaL, there were no withdrawals due to elevated hepatic enzymes.

Hepatic reactions are the most common side effect of Z^9^. The HRZE regimen, in which H and R are also associated with hepatotoxicity,^10^ has served as the control arm in DS-TB in the TB Alliance studies. To understand the relative risks of different regimens used in different trials, in particular the roles of pretomanid and pyrazinamide, we have conducted an exploratory, retrospective analysis of individual-patient data from all the studies conducted by TB Alliance of at least eight weeks duration. The primary objective of the analysis was to compare regimens containing Pa and Z (PaZX), BPaL, and HRZE through eight weeks of treatment in their respective populations. Other aspects of hepatic safety (potential Hy’s Law cases) were also reviewed.

## METHODS

### Data

Data were pooled from all six completed studies conducted by TB Alliance of pretomanid in participants with pulmonary TB where the treatment duration was at least eight weeks: NC-002^3^ (8 weeks; PaMZ versus HRZE in DS-TB, PaMZ in MDR-TB), NC-005^4^ (8 weeks; BPaZ versus HRZE in DS-TB, BPaMZ in MDR-TB), STAND^5^ (4 and 6 months PaMZ versus 6 months HRZE in DS-TB, 6 months PaMZ in MDR-TB), Nix-TB^7^ (6 months; BPaL in highly resistant TB), ZeNix^8^ (6 months; BPaL in highly resistant TB), and SimpliciTB^6^ (4 months BPaMZ versus 6 months HRZE in DS-TB; 6 months BPaMZ in MDR-TB). All research protocols were approved by institutional review boards or ethics committees, and all participants gave written informed consent. Details about ethics committees are provided in the Supplementary Material, as are more details about the study designs and overall results.

Participants included in the analysis were grouped into three sets of regimens: 1) HRZE, 2) PaZ-containing regimens (PaZX), and 3) BPaL. These regimen groups were compared based on treatment-emergent elevations of alanine aminotransferase (ALT), which was selected as an indicator of liver injury because it is considered more specific than aspartate aminotransferase (AST) as well as more objective than adverse event reports^11^.

In the PaZX regimens, pretomanid was dosed as 100 mg or 200 mg once daily (QD), moxifloxacin (M) as 400 mg QD, and pyrazinamide as 1500 mg QD. Bedaquiline was dosed as 400 mg QD for 2 weeks followed by 200 mg thrice weekly in one BPaZ arm of NC-005 and in Nix-TB, or as 200 mg QD for 8 weeks in one BPaZ arm and the BPaMZ arm of NC-005, or as 200 mg QD for 8 weeks followed by 100 mg QD in SimpliciTB and ZeNix. In BPaL, pretomanid was dosed as 200 mg QD. Linezolid was dosed starting at 1200 mg QD in Nix-TB and starting at 600 mg QD or 1200 mg QD in ZeNix; the duration was 26 weeks in Nix-TB and 9 weeks or 26 weeks in ZeNix; in both studies, linezolid dose adjustments for toxicity were allowed. HRZE was dosed by weight bands according to standard practice^12^.

The six studies were conducted at different stages of pretomanid’s development, spanned a decade, and had different schedules of laboratory safety assessments (Table S1); and medical monitoring activities evolved over time. Later protocols had more specific hepatic-safety guidelines, but investigators still had discretion to perform unscheduled visits and to manage participants. All six studies used the DMID toxicity scale (2007 draft^13^; see Supplementary Material), where ALT elevations between >3 and 8 times the upper limit of normal (>3xULN to 8xULN) were classified as grade 3, and ALT elevations more than 8xULN were classified as grade 4. Therefore, elevations >3xULN and >8xULN were the primary focus here, but elevations >5xULN and >10xULN were also examined in alignment with FDA guidance^11^.

All participants who received at least one dose of the relevant study regimens had at least one post-dose ALT measurement, and did not have a baseline value of ALT exceeding the threshold elevation level (3xULN, 5xULN, 8xULN, or 10xULN) were included in the analysis for treatment-emergent exceedance of the given threshold elevation level.

Two studies had only eight weeks of treatment, and in the other four most first occurrences of ALT elevation were by eight weeks. So, here the focus was on the first eight weeks of treatment. For probability calculations, the eight-week timepoint was taken to be Day 60. Other methodological details regarding temporal attribution of events are provided in the Supplemental Material.

### Primary analysis

The three regimen groups’ probabilities of treatment-emergent ALT elevations during the first eight weeks of treatment were estimated using Kaplan-Meier analysis of time to first occurrence that censored participants who withdrew before the end of eight weeks or who did not have an elevation by eight weeks. Also, the probability of an elevation of >3xULN progressing to an elevation >8xULN was estimated by restricting the Kaplan-Meier analysis of the latter event to participants experiencing the former event (and similarly for the progression from >3xULN to >10xULN). All estimated probabilities were compared across the three regimens using log-rank tests.

### Secondary analyses

Two types of secondary analyses were undertaken: 1) Cox regression modelling; 2) qualitative review of other aspects of hepatic safety.

### Cox modeling

Three sets of Cox regression models were applied to assess whether differences among regimen groups identified in the primary analysis remained after accounting for participant characteristics that may vary in distribution across the non-randomized groups.

In the first set, the same three regimen groups of HRZE, PaZX, and BPaL were compared accounting for main effects of baseline xULN, age, weight, sex, race, and HIV status.

The second and third sets assessed the influence of pretomanid pharmacokinetic exposure and DS-TB versus DR-TB. In the second set, only the pretomanid-containing regimens PaZX and BPaL were compared. To adjust for different pretomanid doses and possible pharmacokinetic interactions, steady-state pretomanid exposure, quantified by the 24-hour area under the curve of pretomanid concentration (AUC_0-24_), available from a published population pharmacokinetic model^14^ and unpublished extensions thereof, was also included as a covariate. Additionally, the PaZX group was subdivided into participants with DS-TB (PaZX-DS) or DR-TB (PaZX-DR). All participants on BPaL had DR-TB.

In the third set of models, PaZX-DS and HRZE were compared accounting for main effects of baseline xULN, age, weight, sex, race, and HIV status. HRZE is the standard of care for, and is only used in, DS-TB.

All three sets contained models for ALT elevations >3xULN and >8xULN. Hypotheses of proportional hazards were tested via the R cox.zph function^15^, and remediations guided by Schoenfeld residuals^15^ were applied when significant departures were detected.

### Other aspects of hepatic safety

Treatment-emergent potential Hy’s Law^11^ cases, defined as participants with ALT or AST >3xULN, total bilirubin >2xULN, and alkaline phosphatase <2xULN, were enumerated.

Narratives were provided for deaths in the PaZX group attributed to adverse liver events.

### Significance

For all hypothesis tests (log-rank tests, Cox-model inferences), the threshold of p < 0.05 was used to judge “statistical significance”, even though all analyses were exploratory and *post hoc*, and no adjustments were made for multiplicity.

## RESULTS

### Data

Figure 1 shows the flow of participants from enrollment through inclusion in the analyses based on the four ALT elevation thresholds. Table S2 provides additional information about participant flow through outcomes.

**Figure 1:**
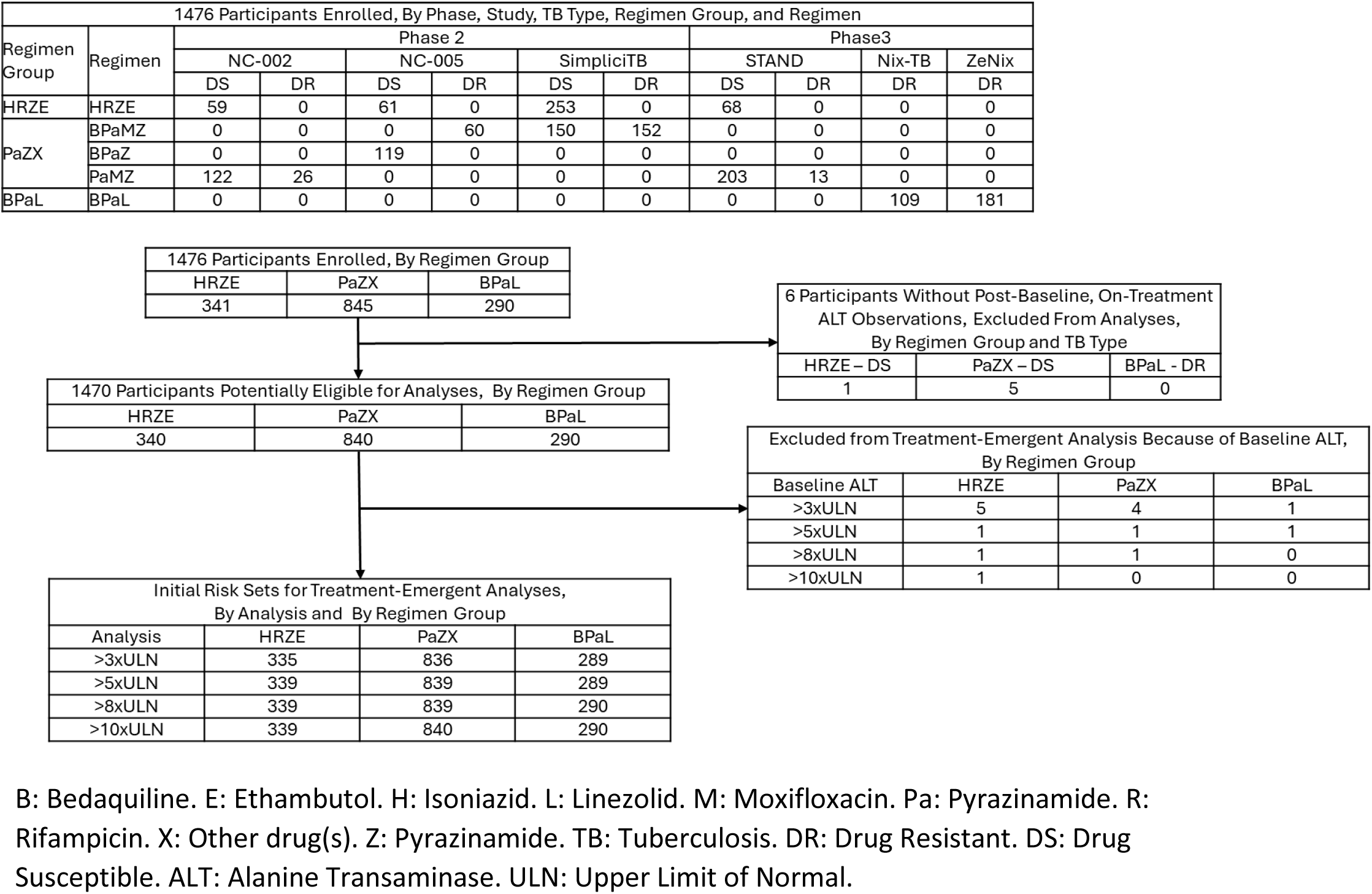
Participant flow from enrollment through risk sets.

Table 1 summarizes patient characteristics. Participants on BPaL were more frequently white or living with HIV. By design, all participants on BPaL had DR-TB and all on HRZE had DS-TB; 70.5% of participants in the PaZX group had DS-TB.

**Table 1:** Table of Summary Statistics of Covariates.

| Variable | PaZX | BPaL | HRZE |
| --- | --- | --- | --- |
| n | 841 | 290 | 340 |
| Baseline xULN: Mean (SD) | 0.593 (0.529) | 0.664 (0.555) | 0.611 (0.801) |
| [N>3xULN; N>8xULN] | [4; 1] | [1; 0] | [5; 1] |
| Age (years): Mean (SD) | 35.1 (12.1) | 36.6 (10.4) | 34.8 (11.5) |
| Weight (kg): Mean (SD) | 55.4 (11.1) | 60.8 (13.7) | 55.0 (10.4) |
| Pretomanid AUC<sub>0-24</sub> (mg.h/L):<br>Mean (SD) | 73.3 (31.0)<sup>1</sup> | 71.6 (31.5)<sup>2</sup> | NA |
| Sex Female: N (%) | 259 (30.8) | 111 (38.3) | 88 (25.9) |
| Race Asian: N (%) | 36 (4.3) | 0 (0.0) | 7 (2.1) |
| Race Black: N (%) | 586 (69.7) | 149 (51.4) | 254 (74.7) |
| Race Other: N (%) | 147 (17.5) | 25 (8.6) | 51 (15.0) |
| Race White: N (%) | 72 (8.6) | 116 (40.0) | 28 (8.2) |
| HIV Positive: N (%) | 185 (22.0) | 92 (31.7) | 67 (19.7) |
| TB Type DS: N (%) | 593 (70.5%) | 0 (0.0%) | 340 (100.0%) |

B: Bedaquiline. E: Ethambutol. H: Isoniazid. L: Linezolid. Pa: Pyrazinamide. R: Rifampicin. X: Other drug(s). Z: Pyrazinamide. ULN: Upper Limit of Normal. SD: Standard Deviation. N: Number kg: kilograms. mg: milligrams. h: hours. L: Liters. HIV: Human Immunodeficiency Virus. TB: Tuberculosis. DS: Drug Susceptible.
<sup>1</sup>n=831. <sup>2</sup>n=277.

### Primary analysis

Table 2, Figure 2, and Figure S1 summarize the primary analysis. PaZX had a significantly greater risk than HRZE for ALT elevations >3xULN, and PaZX had a significantly greater risk than BPaL for ALT elevations >8xULN.

**Figure 2:**
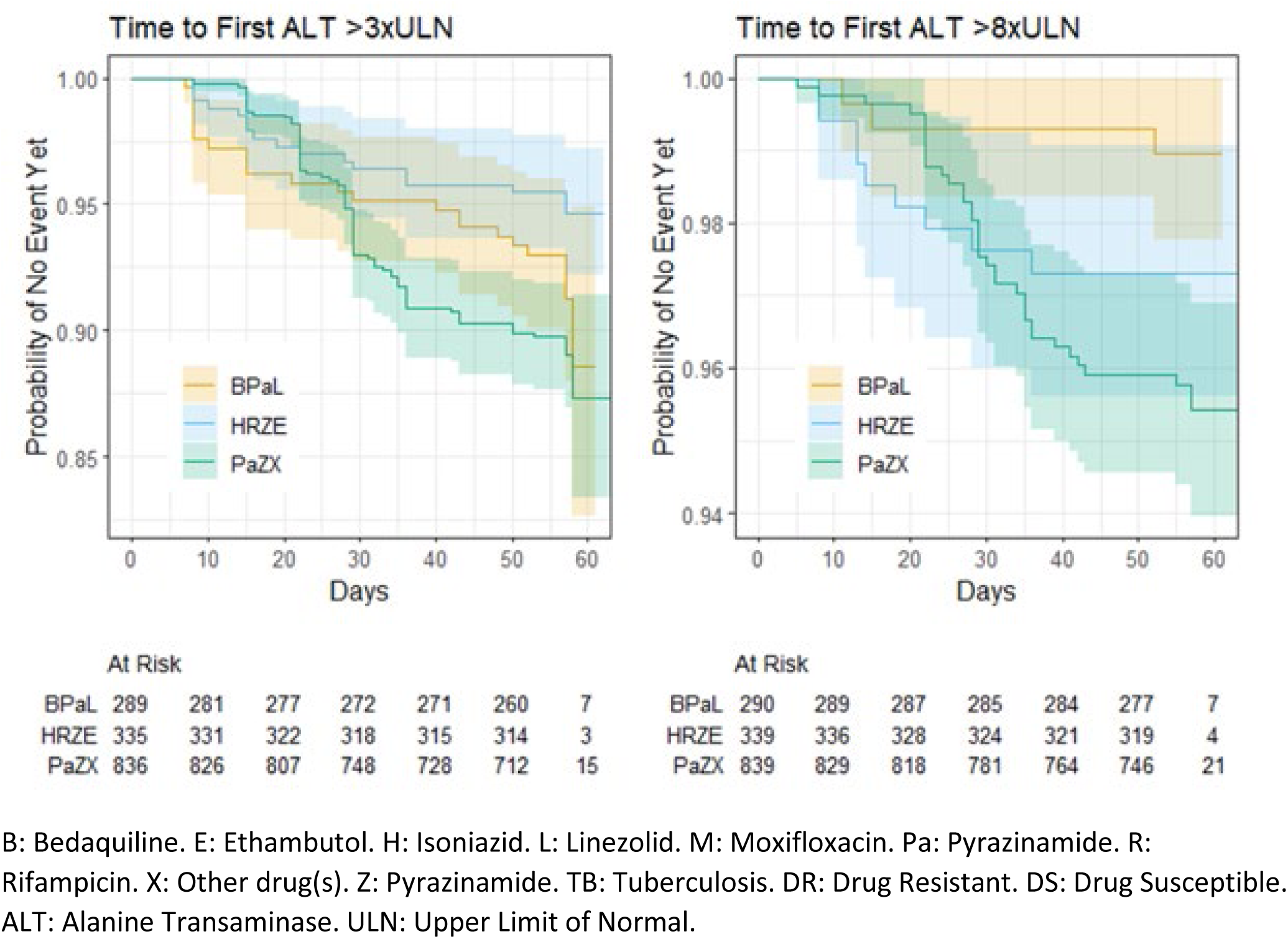
Time to First Elevation of More Than 3xULN and More than 8xULN by Regimen Group.

**Table 2:** Probabilities of Treatment-Emergent ALT Elevation During the First 8 Weeks of Treatment by Different Pooling Groups.

| Statistic | HRZE | PaZX | BPAL | p-values<sup>a</sup> |
| --- | --- | --- | --- | --- |
| P(>3xULN)<sup>b</sup> | 17/335<br>5.36 [2.82-7.82] % | 88/836<br>12.7 [8.59-16.6] % | 25/289<br>11.4 [5.07-17.3] % | 0.011<br>0.003, 0.098, 0.276 |
| P(>5xULN) | 13/339<br>3.87 [1.79-5.92] % | 61/839<br>7.71 [5.82-9.55] % | 9/289<br>3.34 [1.16-5.47] % | 0.006<br>0.027, 0.596, 0.009 |
| P(>8xULN) | 9/339<br>2.68 [0.939-4.40] % | 36/839<br>4.58 [3.10-6.03] % | 3/290<br>1.05 [0-2.23] % | 0.018<br>0.177, 0.136, 0.007 |
| P(>10xULN) | 4/339<br>1.20 [0.0240-2.36] % | 36/840<br>4.57 [3.09-6.02] % | 2/290<br>0.692 [0-1.64] % | 0.001<br>0.007, 0.523, 0.003 |
| P(>8xULN\|>3xULN)<sup>c</sup> | 8/17<br>50.9 [18.0-70.6] % | 35/88<br>45.0 [32.1-55.4] % | 3/25<br>12.0 [0-23.9] % | 0.006<br>0.16, 0.005, 0.007 |
| P(>10xULN\|>3xULN) | 4/17<br>28.7 [0-49.5] % | 34/88<br>43.9 [31.0-54.4] % | 2/25<br>8.00 [0-18.0] % | 0.016<br>0.639, 0.122, 0.004 |

B: Bedaquiline. E: Ethambutol. H: Isoniazid. L: Linezolid. Pa: Pyrazinamide. R: Rifampicin. X: Other drug(s). Z: Pyrazinamide. ULN: Upper Limit of Normal.
<sup>a</sup>p-values are for tests of equal probabilities within a row. The top p-value is for the test of all three probabilities being equal, HRZE=PaZX=BPAL. The bottom three p-values are for HRZE=PaZX, HRZE=BPAL, PaZX=BPAL, respectively.
<sup>b</sup>P(>nxULN): (Number of Participants with Events)/(Number Without ALT > nxULN at Baseline); Estimated probability of Treatment-Emergent ALT > nxULN by Day 60 [95% Confidence Interval], computed by Kaplan-Meier analysis.
<sup>c</sup>P(>nxULN|>3xULN): (Number of Participants with Events)/(Number With Treatment Emergent ALT > 3xULN); Estimated Probability of Treatment-Emergent ALT > nxULN by Day 60, Given Treatment-Emergent >3xULN by Day 60, computed by Kaplan-Meier analysis restricted to participants with Treatment-Emergent ALT >3xULN.

For those who developed an elevation >3xULN, the probability of progressing to >8xULN was significantly lower for BPaL versus either HRZE or PaZX.

Figure 2 shows qualitatively different profiles of time to first elevation, most evident for >8xULN, with the PaZX curve relatively flat through three weeks but then accelerating and crossing the curves for BPaL and PaZX.

### Secondary analyses

#### Cox modeling

Table 3 summarizes results for >3xULN. As in the primary analysis, PaZX was found to have significantly greater risk than HRZE. In addition, here PaZX was found to have significantly greater risk than BPaL.

**Table 3:** Results from Cox Model for Time to First Elevation More than 3xULN, HRZE versus PaZX versus BPaL.

| Variable | Coefficient | Hazard Ratio | Standard Error of Coefficient | p-value |
| --- | --- | --- | --- | --- |
| HRZE:PaZX | -0.688 | 0.502 | 0.266 | 0.010 |
| BPaL:PaZX | -0.568 | 0.567 | 0.235 | 0.016 |
| HRZE:BPaL | -0.121 | 0.886 | 0.322 | 0.708 |
| Baseline | 0.650 | 1.92 | 0.171 | < 0.001 |
| Age | 0.0177 | 1.02 | 0.00752 | 0.019 |
| Weight | 0.0148 | 1.01 | 0.00658 | 0.024 |
| Female | 0.421 | 1.52 | 0.189 | 0.026 |
| Non-Black | 0.491 | 1.63 | 0.206 | 0.017 |
| HIV Positive | 0.453 | 1.57 | 0.221 | 0.041 |

B: Bedaquiline. E: Ethambutol. H: Isoniazid. L: Linezolid. Pa: Pyrazinamide. R: Rifampicin. X: Other drug(s). Z: Pyrazinamide. HIV: Human Immunodeficiency Virus.
From a Cox model with reference values PaZX, Baseline xULN = 0.6, Age = 35 years, Weight = 55 kg, Male, Black, HIV negative. P-values based on standard normal distribution for Coefficient/Standard Error of Coefficient. Inference for HRZE:BPaL derived from estimates and covariance matrix for HRZE:PaZX and BPaL:PaZX.

HRZE and BPaL were not significantly different from each other. Increasing baseline ALT, age, and weight were all associated with increasing risk, as were female, non-Black, and HIV-positive status.

Table S3 shows similar results for >8xULN. PaZX was the reference group, with main effects for HRZE and BPaL. As in the primary analysis, PaZX had significantly greater risk than BPaL. The hypothesis of proportional hazards was rejected for HRZE. Based on Schoenfeld residuals, a piecewise-linear effect was modeled, decreasing to 40 days then flat. The hazard rate was significantly greater than that of PaZX initially but significantly smaller at Day 40. The hazard rate for HRZE was also significantly greater than that of BPaL initially but not later. Covariates had qualitatively similar effects as with >3xULN, but weight and HIV status were not statistically significant. Baseline also required a time-dependent hazard, decreasing piecewise linearly to Day 25.

Tables S4 and S5 show results comparing PaZX-DS, PaZX-DR, and BPaL, with pretomanid AUC as an additional covariate. The hazard rate for PaZX-DS was significantly greater than that of both PaZX-DR and BPaL, and the latter two were not significantly different. Pretomanid AUC did not have a significant effect.

Tables S6 and S7 show results comparing HRZE versus PaZX-DS. For >3xULN, HRZE had a significantly lower hazard rate than PaZX-DS. For >8xULN, the hypothesis of proportional hazards was rejected, and HRZE was found to have a significantly higher hazard rate initially and a significantly lower hazard rate later.

### Other aspects of hepatic safety

There were 3, 7, and 2 treatment-emergent potential Hy’s Law cases in the HRZE, PaZX, and BPaL groups, respectively. The two cases from BPaL had clear alternative etiology. See Table S8 for details.

### Deaths

Narratives of deaths in the PaZX group attributed to adverse liver events are presented in the Supplementary Material.

## DISCUSSION

In this analysis across six studies with almost 1,500 participants, we characterized the hepatic safety profiles of three treatment regimens: HRZE, PaZX, and BPaL. Compared to PaZX, the BPaL group had lower rates of ALT elevations >8xULN.

Hepatotoxicity is an established risk of pyrazinamide^16–19^, so the behavior of PaZX may be due to pyrazinamide. However, ALT elevations were found to be more frequent for PaZX than for HRZE, albeit with HRZE having higher hazard early in treatment, suggesting a contribution of pretomanid or interaction between pretomanid and pyrazinamide. The observation of greater risk for PaZX-DS versus PaZX-DR and the excess hazard for PaZX relative to HRZE later in treatment suggests an immunological mechanism may be involved in the process. BPaL is currently being tested for the first time in a DS-TB population in the NC-009 study (NCT06058299). This will provide an opportunity to determine whether the difference between DS-TB and DR-TB for PaZX applies also to BPaL.

The preclinical program of pretomanid found no evidence of hepatotoxic potential at clinical exposure levels. The potential for pretomanid to cause liver changes was determined in repeat-dose toxicity studies in rodents and monkeys up to 39 weeks duration^20^. Although hepatocellular hypertrophy was seen in mice, rats, and monkeys given daily oral doses of pretomanid at dose levels ≥ 100 mg/kg/day (which is ≥487 mg human-equivalent dose assuming a 60 kg patient^21^), it was considered an adaptive response associated with increased metabolism. The pretomanid plasma exposures at doses where no adverse liver changes occurred ranged from approximately 2- to 5-fold higher than the clinical efficacious exposure.

Pretomanid monotherapy for up to 2 weeks was well tolerated in healthy participants^22–24^ and participants with TB^25,26^. Healthy participants received pretomanid 50 mg – 1500 mg single dose and 200 mg – 1000 mg multiple dose for 7 or 8 days. One healthy participant (1/289, 0.35%) on pretomanid multiple dose 200 mg had a treatment-emergent ALT elevation that was between 3xULN and 8xULN. Participants with DS-TB received 50 – 1200 mg pretomanid for 14 days in two early bactericidal activity studies. One participant (1/122, 0.82%) on pretomanid multiple dose 1200 mg had a treatment-emergent ALT elevation between 3xULN and 8xULN.

This exploratory, retrospective analysis has several limitations. Samples sizes were small relative to the rarity of the events of interest, reducing precision and power. There were imbalances in participant characteristics, exposures, and pyrazinamide doses between regimens in the cross-study comparisons. The studies were conducted over the span of a decade, and changes in patient care for TB, HIV, and other comorbidities over this period could have influenced observations. Study designs and medical monitoring activities also evolved with time and experience. Although guidelines were provided for handling hepatoxicity, investigators could decide how to manage participants, potentially introducing additional heterogeneity. Moreover, the studies were open-label or partially so, which could have influenced investigators’ decisions. Geography might be another important factor, as different countries may have different practices; some of the trials were conducted only in one or a few countries while other trials had broader footprints. Other limitations include different frequencies of laboratory testing in the first 8 weeks, and incomplete information to systematically assess concurrent risk factors of liver injury such as co-infection with viral hepatitis, concomitant medications, alcohol use, and malnutrition.

## CONCLUSION

ALT elevations that occurred in different pretomanid-containing regimens had different characteristics. Participants receiving PaZ-containing regimens had a higher incidence of elevations >8xULN compared with participants receiving BPaL. The hepatic safety profile of BPaL in its indicated DR-TB population has been generally similar to that of HRZE in its indicated DS-TB population. Pretomanid and pyrazinamide should be co-administered only when the benefit outweighs the risk.

## Author contributions

Conception and design of the study: ML, JN

Acquisition of data: ML, MO, MC, SG, DS Analysis and interpretation of data: JN, ML

Participation in drafting or revision of the submitted article: JN, ML, MO, RB-B, JA, MC, SG, DS, MB, ES

## Data availability

Data are or will soon be available at TB-PACTS (TB-PACTS | Critical Path Institute (c-path.org)). As described in the Narrative for Case 3 and in Table S11, ALT elevations for one participant were hard-coded from local labs and are not included in the data at TB-PACTS.

## SUPPLEMENTARY MATERIAL

### Descriptions of studies

#### NC-002

Title: A Phase 2 Open-Label Partially Randomized Trial to Evaluate the Efficacy, Safety, and Tolerability of the Combination of Moxifloxacin plus PA-824 plus Pyrazinamide after 8 weeks of Treatment in Adult Patients with Newly Diagnosed Drug-Sensitive or Multidrug-Resistant, Smear-Positive Pulmonary Tuberculosis^1^

Dates: First-patient-first-visit 20 March 2012 – Last-patient-last-visit 26 July 2013 Countries: Brazil, South Africa, Tanzania

Arms:

- Randomized:

`o` DS-TB: Pa_100_M_400_Z_1500_ once daily for 8 weeks (n = 60)
`o` DS-TB: Pa_200_M_400_Z_1500_ once daily for 8 weeks (n = 62)
`o` DS-TB: HRZE for 8 weeks (n = 59)
- Non-randomized:

`o` MDR-TB: Pa_200_M_400_Z_1500_ once daily for 8 weeks (n = 26)

From Reference 1: “In patients with drug-susceptible tuberculosis, the bactericidal activity of MPa_200_Z (n=54) on days 0–56 (0·155, 95% Bayesian credibility interval 0·133–0·178) was significantly greater than for HRZE (n=54, 0·112, 0·093–0·131). DR-MPa_200_Z (n=9) had bactericidal activity of 0·117 (0·070–0·174). The bactericidal activity on days 7–14 was strongly associated with bactericidal activity on days 7–56.

Frequencies of adverse events were similar to standard treatment in all groups. The most common adverse event was hyperuricaemia in 59 (29%) patients (17 [28%] patients in MPa_100_Z group, 17 [27%] patients in MPa_200_Z group, 17 [29%] patients in HRZE group, and 8 [31%] patients in DR-MPa_200_Z group). Other common adverse events were nausea in (14 [23%] patients in MPa_100_Z group, 8 [13%] patients in MPa_200_Z group, 7 [12%] patients in HRZE group, and 8 [31%] patients in DR-MPa_200_Z group) and vomiting

(7 [12%] patients in MPa_100_Z group, 7 [11%] patients in MPa_200_Z group, 7 [12%] patients in HRZE group, and 4 [15%] patients in DR-MPa_200_Z group). No on-treatment electrocardiogram occurrences of corrected QT interval more than 500 ms (an indicator of potential of ventricular tachyarrhythmia) were reported. No phenotypic resistance developed to any of the drugs in the regimen.”

#### NC-005

Title: A Phase 2 Open-Label Partially Randomized Trial to Evaluate the Efficacy, Safety and Tolerability of Combinations of Bedaquiline, Moxifloxacin, PA-824 and Pyrazinamide During 8 Weeks of Treatment in Adult Subjects with Newly Diagnosed Drug-Sensitive or Multi Drug-Resistant, Smear-Positive Pulmonary Tuberculosis^2^

Dates: First-patient-first-visit 23 October 2014 – Last-patient-last-visit 23 January 2018 Countries: South Africa, Tanzania, Uganda

Arms:

- Randomized:

`o` DS-TB: B_load_Pa_200_Z_1500_ for 8 weeks^1^ (n = 59^2^)
`o` DS-TB: B_200_Pa_100_Z_1500_ once daily for 8 weeks (n = 60)
`o` DS-TB: HRZE for 8 weeks (n = 61)
- Not randomized:

`o` MDR-TB: B_200_Pa_200_M_400_Z_1500_ once daily for 8 weeks (n = 60)

^1^B_load_ = 400 mg once daily for 2 weeks then 200 mg thrice weekly for 6 weeks. Pa_200_ and Z_1500_ once daily for 8 weeks.

^2^One participant with no post-baseline ALT measurements was excluded from the analysis here, hence the sample size for BPaZ in Table 1 is 118, not 119.

From Reference 2: “57 patients in the B_load_PaZ group, 56 in the B_200_PaZ group, and 59 in the HRZE group were included in the primary analysis. B_200_PaZ produced the highest daily percentage change in TTP (5·17% [95% Bayesian credibility interval 4·61–5·77]), followed by B_load_PaZ (4·87% [4·31–5·47]) and HRZE group (4·04% [3·67–4·42]). The bactericidal activity in B_200_PaZ and B_load_PaZ groups versus that in the HRZE group was significantly different. Higher proportions of patients in the B_load_PaZ (six [10%] of 59) and B_200_PaZ (five [8%] of 60) groups discontinued the study drug than in the HRZE group (two [3%] of 61) because of adverse events. Liver enzyme elevations were the most common grade 3 or 4 adverse events and resulted in the withdrawal of ten patients (five [8%] in the B_load_PaZ group, three [5%] in the B_200_PaZ group, and two [3%] in the HRZE group). Serious treatment-related adverse events affected two (3%) patients in the B_load_PaZ group and one (2%) patient in the HRZE group. Seven (4%) patients with drug-susceptible tuberculosis died and four (7%) patients with rifampicin-resistant tuberculosis died. None of the deaths were considered to be related to treatment.”

#### STAND

Title: A Phase 3 Open-Label Partially Randomized Trial to Evaluate the Efficacy, Safety and Tolerability of the Combination of Moxifloxacin plus PA-824 plus Pyrazinamide after 4 and 6 months of Treatment in Adult Subjects with Drug-Sensitive Smear-Positive Pulmonary Tuberculosis and after 6 months of Treatment in Adult Subjects with Multi-Drug Resistant, Smear-Positive Pulmonary Tuberculosis^3^

Dates: First-patient-first-visit 19 February 2015 – Last-patient-last-visit 29 November 2017 Countries: Georgia, Kenya, Malaysia, Philippines, South Africa, Thailand, Uganda, Ukraine, Tanzania Arms:

- Randomized:

`o` DS-TB: Pa_100_M_400_Z_1500_ once daily for 4 months (n = 65)
`o` DS-TB: Pa_200_M_400_Z_1500_ once daily for 4 months (n = 71)
`o` DS-TB: Pa_200_M_400_Z_1500_ once daily for 6 months (n = 67)
`o` DS-TB: HRZE for 2 months/HR for 4 months (n = 68)
- Not randomized:

`o` MDR-TB: Pa_200_M_400_Z_1500_ once daily for 6 months (n = 13)

From Reference 3: “Respectively 4/47 (8.5%), 11/57 (19.3%), 14/52 (26.9%) and 1/53 (1.9%) DS-TB outcomes were unfavourable in patients on 6Pa_200_MZ, 4Pa_200_MZ, 4Pa_100_MZ and controls. There was a 6.6% (95% CI –2.2% to 15.4%) difference per protocol and 9.9% (95%CI –4.1% to 23.9%) modified intention-to-treat difference in unfavourable responses between the control and 6Pa_200_MZ arms. Grade 3 adverse events affected 68/203 (33.5%) receiving experimental regimens, and 19/68 (27.9%) on control. Ten of 203 (4.9%) participants on experimental arms and 2/68 (2.9%) controls died.”

#### Nix-TB

Title: A Phase 3, Open-label Trial Assessing the Safety and Efficacy of Bedaquiline Plus Pretomanid Plus Linezolid in Subjects with Pulmonary Infection of Either Extensively Drug-resistant Tuberculosis (XDR-TB) or Treatment Intolerant/Non-responsive Multi-Drug Resistant ^Tuberculosis^ (MDR-TB)^4^

Dates: First-patient-first-visit 16 April 2015 – Last-patient-last-visit 3 August 2020 Country: South Africa

Arms:

- Not randomized:

`o` DR-TB^1^:B_load_Pa_200_L_1200_ for 26 weeks^2,3^ (n = 109)

^1^See Title for types of DR-TB.

^2^B_load_ = 400 mg once daily for 2 weeks then 200 mg thrice weekly for 24 weeks. Pa_200_ once daily for 26 weeks.

^3^L1200 initially as once daily then amended to 600 mg twice daily.

From Reference 4: “At 6 months after the end of treatment in the intention-to-treat analysis, 11 patients (10%) had an unfavorable outcome and 98 patients (90%; 95% confidence interval, 83 to 95) had a favorable outcome. The 11 unfavorable outcomes were 7 deaths (6 during treatment and 1 from an unknown cause during follow-up), 1 withdrawal of consent during treatment, 2 relapses during follow-up, and 1 loss to follow-up. The expected linezolid toxic effects of peripheral neuropathy (occurring in 81% of patients) and myelosuppression (48%), although common, were manageable, often leading to dose reductions or interruptions in treatment with linezolid.”

### ZeNix

Title: A Phase 3 Partially-blinded, Randomized Trial Assessing the Safety and Efficacy of Various Doses and Treatment Durations of Linezolid Plus Bedaquiline and Pretomanid in Participants with Pulmonary Infection of Either Extensively Drug-resistant Tuberculosis (XDR-TB), Pre-XDR-TB or Treatment Intolerant or Non-responsive Multi-drug Resistant Tuberculosis (MDR-TB)^5^

Dates: First-patient-first-visit 16 November 2017 – Last-patient-last-visit 26 November 2021 Countries: Georgia, Moldova, Russia, South Africa

Arms:

- Randomized:

`o` DR-TB^1^: Linezolid 1200 mg for 26 weeks: B_QD_Pa_200_L_1200_ once daily for 26 weeks^2^ (n = 45)
`o` DR-TB: Linezolid 1200 mg for 9 weeks: B_QD_Pa_200_ once daily for 26 weeks plus L_1200_ once daily for 9 weeks.
`o` DR-TB: Linezolid 600 mg for 26 weeks: B_QD_Pa_200_L_600_ once daily for 26 weeks (n = 45)
`o` DR-TB: Linezolid 600 mg for 9 weeks: B_QD_Pa_200_ once daily for 26 weeks plus L_600_ once daily for 9 weeks. (n = 45)

^1^See Title for types of DR-TB.

^2^BQD = 200 mg once daily for 8 weeks then 100 mg once daily for 18 weeks.

From Reference 5: “Among participants who received bedaquiline–pretomanid–linezolid with linezolid at a dose of 1200 mg for 26 weeks or 9 weeks or 600 mg for 26 weeks or 9 weeks, 93%, 89%, 91%, and 84%, respectively, had a favorable outcome; peripheral neuropathy occurred in 38%, 24%, 24%, and 13%, respectively; myelosuppression occurred in 22%, 15%, 2%, and 7%, respectively; and the linezolid dose was modified (i.e., interrupted, reduced, or discontinued) in 51%, 30%, 13%, and 13%, respectively. Optic neuropathy developed in 4 participants (9%) who had received linezolid at a dose of 1200 mg for 26 weeks; all the cases resolved. Six of the seven unfavorable microbiologic outcomes through 78 weeks of follow-up occurred in participants assigned to the 9-week linezolid groups.”

#### SimpliciTB

Title: An Open-Label, Partially Randomized Trial to Evaluate the Efficacy, Safety and Tolerability of a 4-month Treatment of Bedaquiline plus Pretomanid plus Moxifloxacin plus Pyrazinamide (BPaMZ) Compared to a 6-month Treatment of HRZE/HR (Control) in Adult Participants with Drug-Sensitive Smear-Positive Pulmonary Tuberculosis (DS-TB) and a 6-month Treatment of BPaMZ in Adult Participants with Drug Resistant, Smear-Positive Pulmonary Tuberculosis (DR-TB)^6^

Dates: First-patient-first-visit 23 July 2018 – Last-patient-last-visit 5 April 2022 Countries: Brazil, Georgia, Phillipines, Russia, South Africa, Tanzania, Uganda Arms:

- Randomized:

`o` DS-TB: B_QD_Pa_200_M_400_Z_1500_ once daily for 17 weeks (n = 150)
`o` DS-TB: HRZE for 2 months/HR for 4 months (n = 153^1^)
- Not randomized:

`o` DR-TB^2^: B_QD_Pa_200_M_400_Z_1500_ once daily for 26 weeks (n = 152^3^)

^1^One participant with no post-baseline ALT measurements was excluded from the analysis here, hence the sample size for HRZE in Table 1 is 152, not 153.

^2^Participants with DR-TB were defined as mono-resistance to rifampicin or isoniazid, or resistance to both rifampicin and isoniazid (MDR-TB).

^3^Three participants with no post-baseline ALT measurements were excluded from the analysis here, hence the sample size for BPaMZ in Table 1 is 299, not 302.

From Reference 6: “In a modified intention to treat analysis (mITT), by week 8, 122 (84.1%) and 70 (47.3%) participants respectively were culture negative on 4BPaMZ and HRZE with a Hazard Ratio for earlier negative status of 2.93 (95% CI, 2.17-3.96). Median time to negative culture was 6 weeks (IQR: 4-8) on 4BPaMZ and 11 weeks (IQR: 6-12) on HRZE. 152 DR-TB participants received 6BPaMZ with a median time to negative culture of 5 weeks (IQR 3-7). At week 52, 17%, 7% and 17% of participants had unfavourable outcomes on 4BPaMZ, HRZE and 6BPaMZ, respectively. The absolute difference between DS-TB arms was 9.72% (95% CI, 2.35-17.09%) so 4BPaMZ did not meet the non-inferiority margin of 12% in the microbiologically eligible and assessable analysis population (TB-mITT). Higher unfavourable outcome rates on 4BPaMZ were due to withdrawals for adverse events, predominantly related to elevated hepatic enzymes which precluded treatment completion in 7% of participants.”

## Additional methodological details

### Introduction

The objective of this investigation was to compare three regimen groups – BPaL, PaZX, and HRZE – with respect to the probability of hepatotoxicity over eight weeks of treatment using time-to-event (TTE) analysis.

Data were pooled from six studies conducted by TB Alliance. In chronological order, these studies were: NC-002, NC-005, NC-006 = STAND, Nix-TB, NC-007 = ZeNix, and NC-008 = SimpliciTB.

The duration of treatment was eight weeks in NC-002 and NC-005. In the other studies, the duration of treatment was 17 – 26 weeks.

Hepatotoxicity was identified by elevations of ALT to >3xULN, >5xULN, >8xULN, or >10xULN; and, for each value of xULN, the event of the TTE analysis was either the first occurrence of such an elevation of ALT within eight weeks or censoring.

ALT was measured only when participants visited the clinic. Thus, times to event were essentially bound to the times of clinic visits, which generally occurred at intervals of one or two weeks. All studies had visits scheduled immediately after completing the eighth week of treatment. In NC-002 and NC-005, this Week 8 visit was scheduled to occur on Study Day 57, the morning after the 56^th^ treatment day, and was called the Day 57 visit. In the later, longer studies, the visit was referred to as the Week 8 visit, and a day for it was not specified in the protocol. Here, the term *Week 8 visit* will be used for all studies.

However, not all participants in all studies had a Week 8 visit. Some ended treatment before the Week 8 visit (e.g., withdrew, were withdrawn, were lost to follow-up, died, etc.). Others simply missed the visit, especially in the longer studies, where they may have continued treatment through and beyond Week 8 without providing any ALT data at Week 8.

Participants who did have a Week 8 visit did not always make it to the clinic on Day 57 for that visit, even in NC-002 and NC-005 where that was the nominal intent. Thus, the duration of follow-up through eight weeks varied among participants who had treatment through eight weeks.

As stated above, the time span of interest here was eight weeks *of treatment*. Treatment may have stopped before eight weeks, and may not have stopped on a visit day, whereas events could operationally occur only on visit days. The effect, if any, of a treatment on the liver may be expected to extend beyond the exact day of treatment cessation.

Thus, conventions were needed for how to handle missed Week 8 visits, variation in follow-up time through eight weeks, and what it means to be “on treatment” for the purpose of liver toxicity assessment. Here we describe the conventions used.

### Conventions

#### Missing Week 8 visit

A subject who had an event at a visit before Week 8 was counted as such, was removed from the risk set for all later event times, and thus caused no problems.

But for a subject who had no event through the latest visit prior to Week 8 and then continued treatment but did not have a Week 8 visit, it was uncertain whether they should be counted in the risk set for events at Week 8. Therefore, adjustments to the data were made according to the following decision rule:

- Day 50 is identified as the boundary between “before Week 8” and “on or after Week 8”. That is, absent a Week 8 visit, a visit occurring on Day ≤ 50 was considered before Week 8, and a visit occurring on a Day > 50 was considered on or after Week 8.
- If the next visit after Day 50 occurred on or before Day 63, that visit was identified as the terminal visit for the Week 8 analysis, and the subject was considered to have completed through Week 8.
- Otherwise, the latest visit up to and including Day 50 was identified as the terminal visit for the Week 8 analysis, and the subject was considered not to have completed through Week 8 (for the purposes of the Week 8 analysis).
- In either case, if the subject had no event through the terminal visit for the Week 8 analysis, they were censored at that visit for the Week 8 analysis.

#### Variation in “eight weeks”

All visits recognized formally in the source data as Week 8 visits occurred on or after Day 51.

There were 11 subjects for whom visits recognized formally in the source data as Week 8 visits occurred later than Day 61. These were Days 64, 65, 66, 67, 68, 71, 71, 85, 87, 87, and 93.

Thus, only 11 of 1470 subjects in the data pool had Week 8 visits outside of 51 – 61 days.

The existing variability was simply accepted as is. The duration of interest was referred to as “eight weeks” without elaboration.

#### On Treatment

A visit day was considered On Treatment if it occurred no later than seven days after the recorded Treatment End Day.

### Narratives of deaths in the PaZX group attributed to adverse liver events

#### Case 1

Female, 41 – 50 years old, HIV positive, DS-TB, STAND, Pa_200_MZ. Illness Onset: Day 23. Death Day: 28. No other hepatotoxins identified. No autopsy performed.

Initial symptoms were nausea and vomiting, managed with proton pump inhibitor and IV fluid replacement. On Day 25, hyperkalaemia treated with IV calcium gluconate and IV actrapid insulin & 50% dextrose. Patient stopped taking IMP as vomiting. On Day 27, ALT: 7124 U/L, total bilirubin: 168 μmol/L. (These results returned from the lab after death.) No recorded treatment pauses before halting.

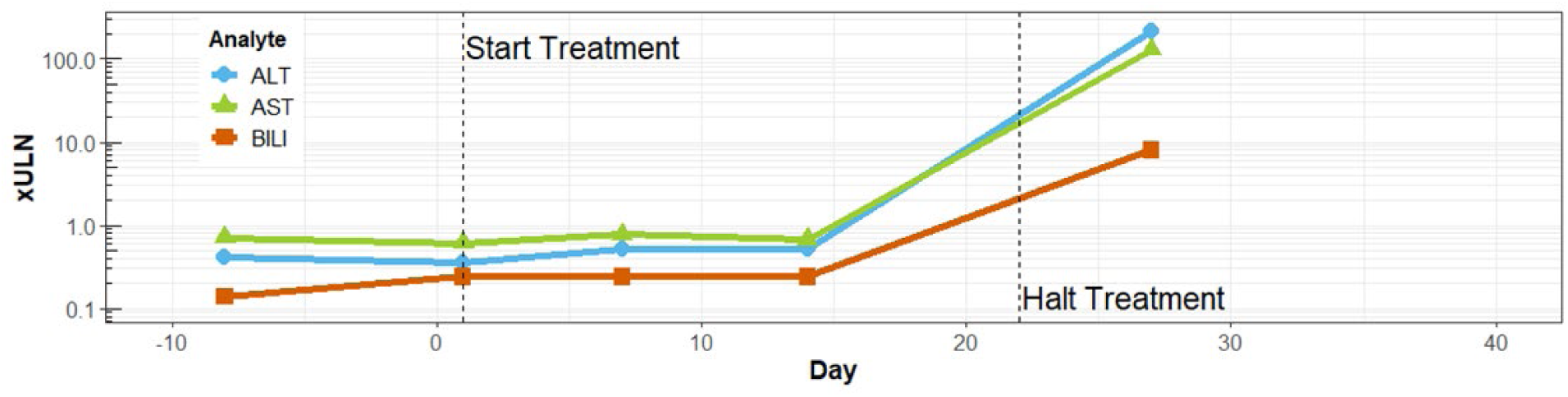

### Case 2

Male, 21 – 30 years old, HIV negative, DS-TB, STAND, Pa_100_MZ. Illness Onset: Day 21. Death Day: 39. No other hepatotoxins identified. Autopsy performed^1^.

Initial symptoms of gastroenteritis, managed supportively without antibiotics. Renal and hepatic biochemistry normal on Day 22 but on Day 29 ALT: 1393 U/L, total bilirubin: 26 μmol/L, and on Day 34 ALT: 3615 U/L, total bilirubin: 185 μmol/L, K: 6.8 mmol/L, creatinine: 206 μmol/L. Patient hospitalised and study medication stopped. Hyperkalaemia treated with IV actrapid insulin & 50% dextrose. IV fluid replacement initiated. Confusion, attributed to hepatic encephalopathy developed after hospitalisation, treated with lactulose. Nutritional support was provided by a dietician. Severe coagulopathy (prothrombin time >120 seconds) was noted on Day 35. Fresh frozen plasma and Vitamin K were administered, alongside a proton pump inhibitor (pantoprazole) and antibiotics (metronidazole). Despite best supportive care, the patient continued to gradually deteriorate with no improvement in the encephalopathy or coagulopathy, both of which were attributed to acute liver failure. No recorded treatment pauses before halting.

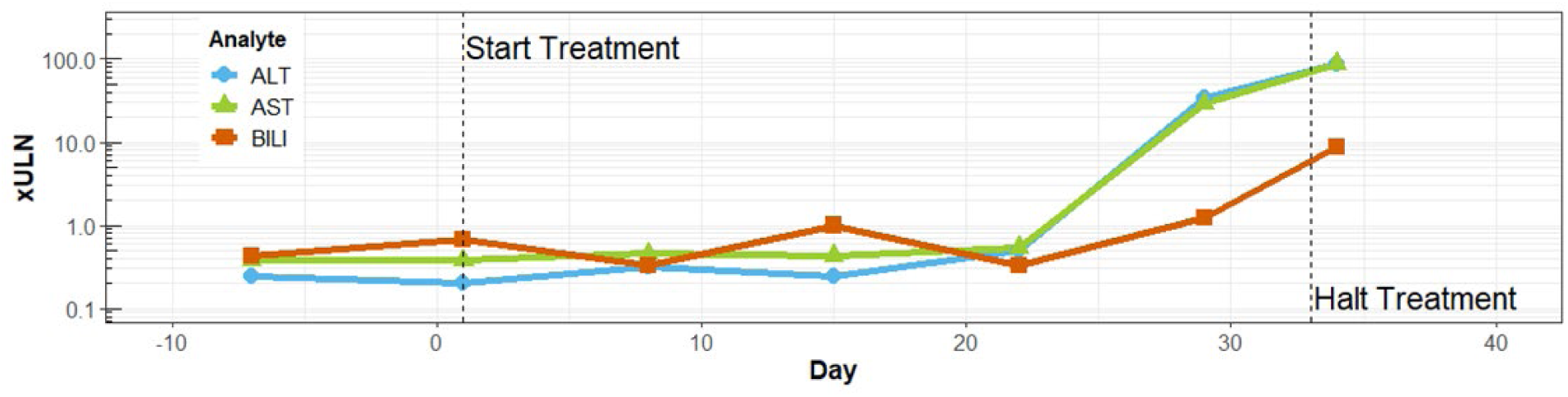

### Case 3

Female, 21 – 30 years old, HIV positive, DS-TB, STAND, Pa_200_MZ. Illness Onset: Day 14. Death Day: 34. No other hepatotoxins identified. No autopsy performed.

Chronic anaemia at baseline (Hb: 6.9 g/dL on screening bloods). Vomiting on Day 14 and 21 but settled each time and felt otherwise well. Hospitalised from Day 25-29 (trial center not informed). On Day 25, ALT: 1649 U/L, total bilirubin: 31 μmol/L, Hb: 6.7 g/dL. Hospital administered transfusion for anaemia. She then developed a rash a few days subsequently that was diagnosed as a transfusion reaction. Re-admitted on Day 32, with confusion, rash, and jaundice. ALT: 2053 U/L, total bilirubin: 171 μmol/L. A formal decision to discontinue all trial medication was taken, and supportive care was given including IV antibiotics (ceftriaxone) and lactulose to manage hepatic encephalopathy. The patient progressively deteriorated and died. Lab values on Days 25 and 32 were obtained from local laboratory and hard-coded into the analysis dataset. No recorded treatment pauses before halting.

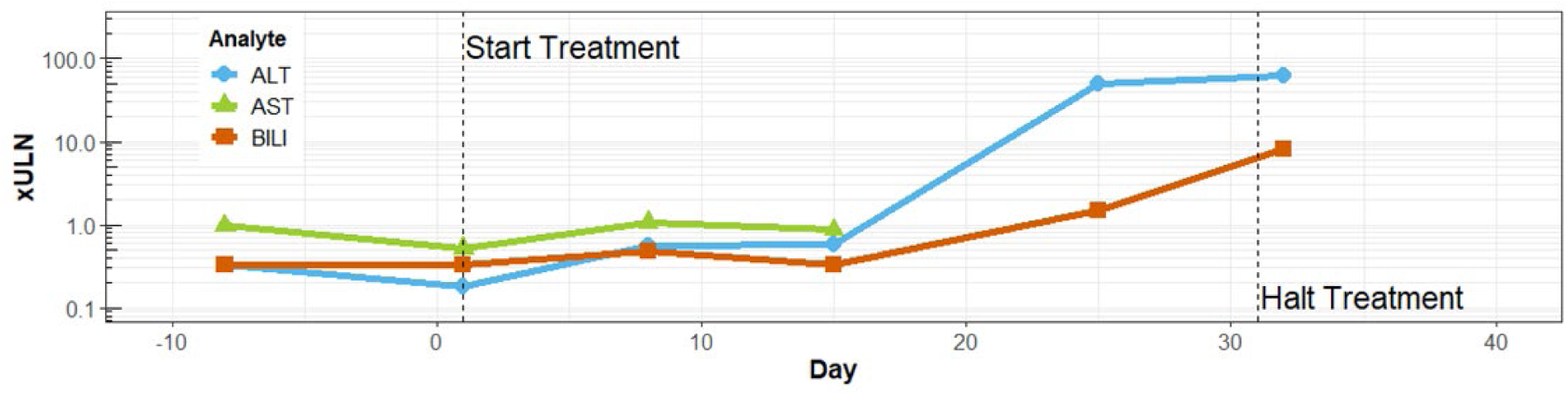

### Case 4

Male, 51 – 60 years old, HIV negative, DS-TB, SimplicTB, BPa_200_MZ. Illness Onset: Day 28. Death Day: 48. Other hepatotoxins: Alcohol, including locally brewed gongo^2^. Autopsy performed^3^.

On Day 28, asymptomatic ALT and AST elevation were noted (71 U/L and 174 U/L). Total bilirubin was 7 μmol/L. On Day 31, ALT: 106 U/L, AST: 243 U/L, total bilirubin: 9 μmol/L. Viral hepatitis serology indicated prior hepatitis A (IgG positive) but no evidence of Hepatitis B or C. On Day 33, he had abdominal pain and diarrhoea. ALT: 321 U/L, AST: 1228 U/L, total bilirubin: 7 μmol/L. All study drugs were stopped. On Day 37, ALT: 948 U/L, AST: 2229 U/L, total bilirubin: 112 umol/L. Thrombocytopenia (platelets: 85x109 cells/L). He was hospitalised and treated supportively, including IV fluids and antibiotics (co-amoxiclav). He developed reduced consciousness, coagulopathy, and deranged kidney function. Despite transfer to the Intensive Therapy Unit for mechanical ventilation, inotropic support, change of antibiotics to IV meropenem, transfusion of fresh frozen plasma and administration of vitamin K, his deterioration continued. No recorded treatment pauses before halting.

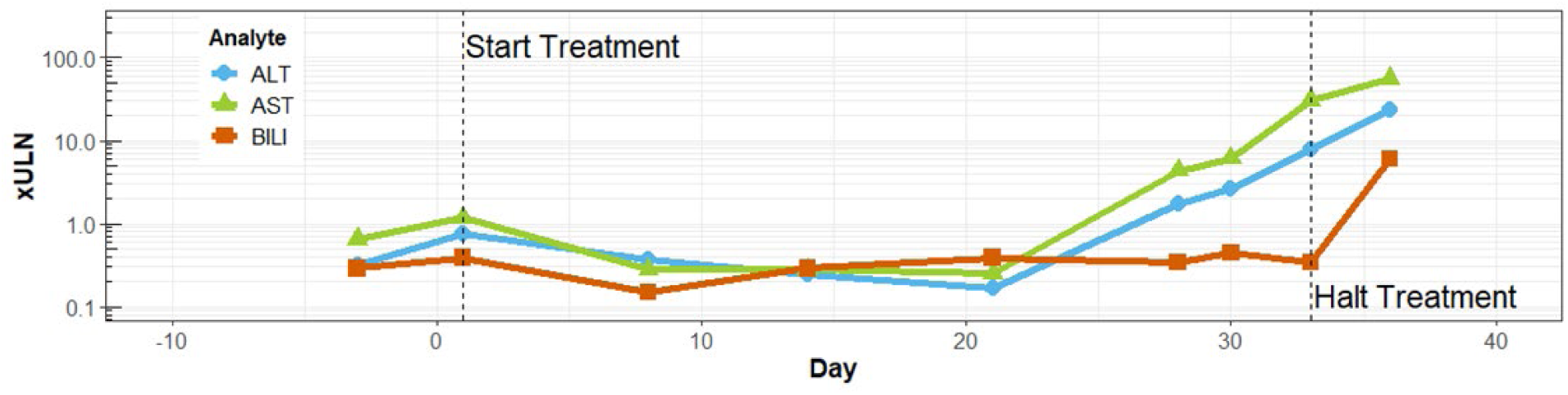

^1^Summary of autopsy result: Tissue sampling from the liver showed extensive autolysis, with no features of granulomatous inflammation, or mass lesions. There was no evidence of interstitial or neoplastic infiltrates, and no vasculitis. Thoracic sections showed left sided pleural effusion, pulmonary oedema, and pneumonia. Kidney samples showed extensive infarction of renal tubules and congestion of renal blood vessels.

^2^Gongo is a highly concentrated, informally brewed alcoholic drink that can contain many impurities including aflatoxin B1 and mycotoxins produced by *Aspergillus flavus* and *Aspergillus parasiticus*. The patient initially denied any consumption of alcoholic drinks but, at a later date, family members and other contacts confirmed that he was repeatedly seen at a local club where Gongo was sold, and his consumption may have been denied because the product was illegal.

^3^Summary of autopsy result: Gross examination of the liver revealed hepatomegaly, micro nodules on the liver, yellowish in color with ascities [ascites]; yellowish fluid. Histologic examination showed fatty liver with distractions of hepatocytes. Gross examination of the kidneys grossly showed “no obvious abnormalities other than mild kidney enlargement.” Renal pathology showed necrosis of tubular lining with cells desquamation and dilated proximal tubule, with acute tubular necrosis and marked glomerular necrosis. Toxicology results indicated the presence of metronidazole in liver, kidney, spleen, lung, and blood samples, consistent with treatment the participant received during hospitalization.

Blood alcohol test was negative. A second pathologist was asked to review these findings but requested additional information which could not be provided. An independent consultant hepatologist also reviewed this case, noting that: “The most important element… is not chronic disease but acute liver failure, evident by the clinical picture and supported by the findings of both pathologists. Overall, it seems likely that this is acute-on-chronic liver injury resulting in death from liver failure and terminal sepsis. The probable cause is one or more drugs in the pretomanid/bedaquiline/moxifloxacin/pyrazinamide regimen, and a contribution of alcohol injury and even aflatoxin hepatotoxicity cannot be excluded, based on available evidence.”

## Figures

**Figure S1:**
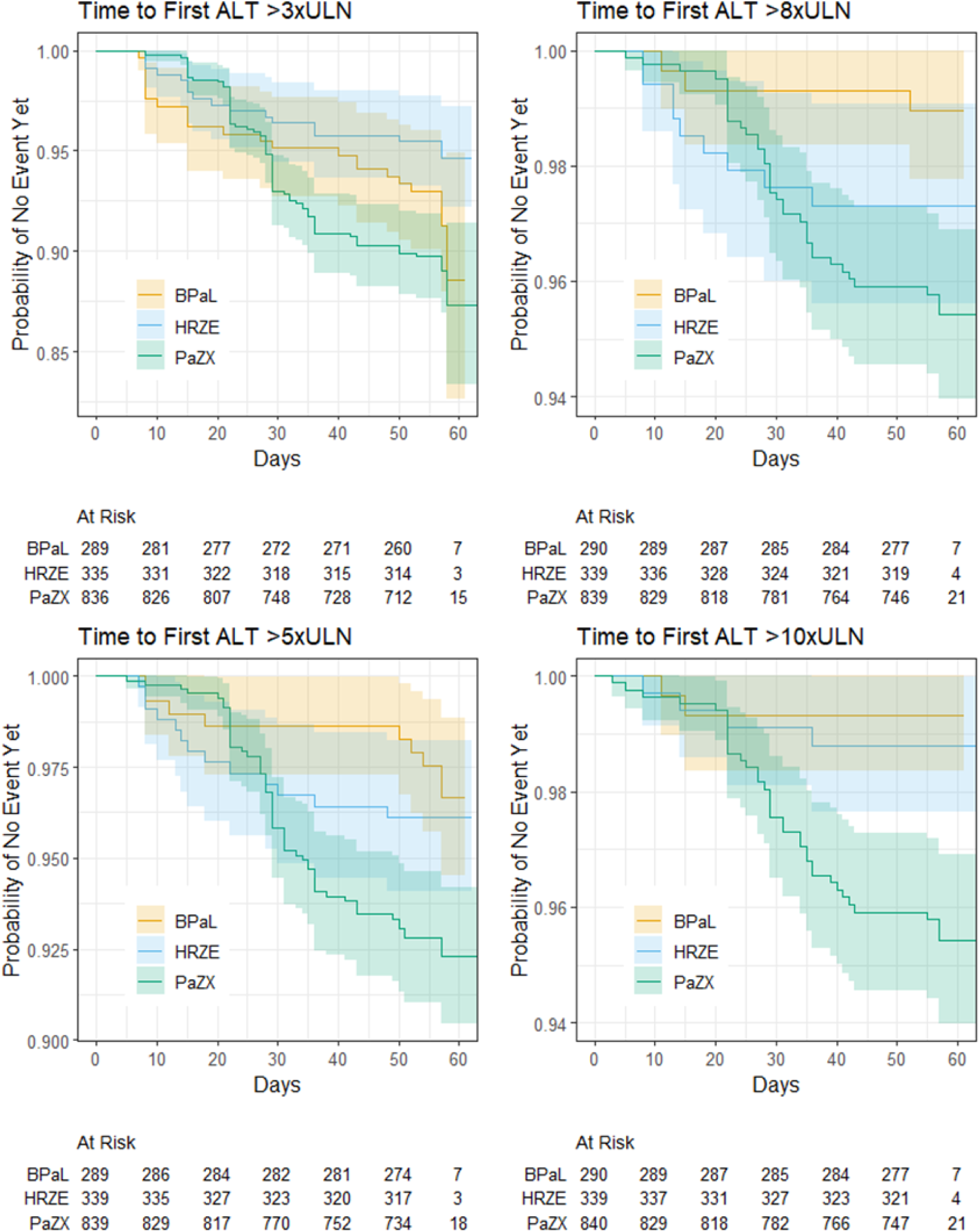
Time to First Elevation of More Than 3xULN / 5xULN / 8xULN / 10xULN by Regimen Group.

## Tables

**Table S1:** Scheduled Visits for Laboratory Safety Tests Per Protocol Post Randomization.

| TB Alliance completed studies (treatment ≥8 weeks) |  | Day 1 | Week 1 | Week 2 | Week 3 | Week 4 | Week 5 | Week 6 | Week 7 | Week 8 |
| --- | --- | --- | --- | --- | --- | --- | --- | --- | --- | --- |
| Phase 2 | NC-002 | X | X | X | X | X | X | X | X | X |
|  | NC-005 | X | X | X | X | X | X | X | X | X |
|  | SimpliciTB | X | X | X | X | X | X | X | X | X |
| Phase 3 | NC-006 | X | X | X |  | X |  |  |  | X |
|  | Nix-TB | X | X | X |  | X |  | X |  | X |
|  | ZeNix | X | X | X | X | X |  | X |  | X |

Note: Investigators had the discretion to request unscheduled visit at any time during the study conduct.

**Table S2:**
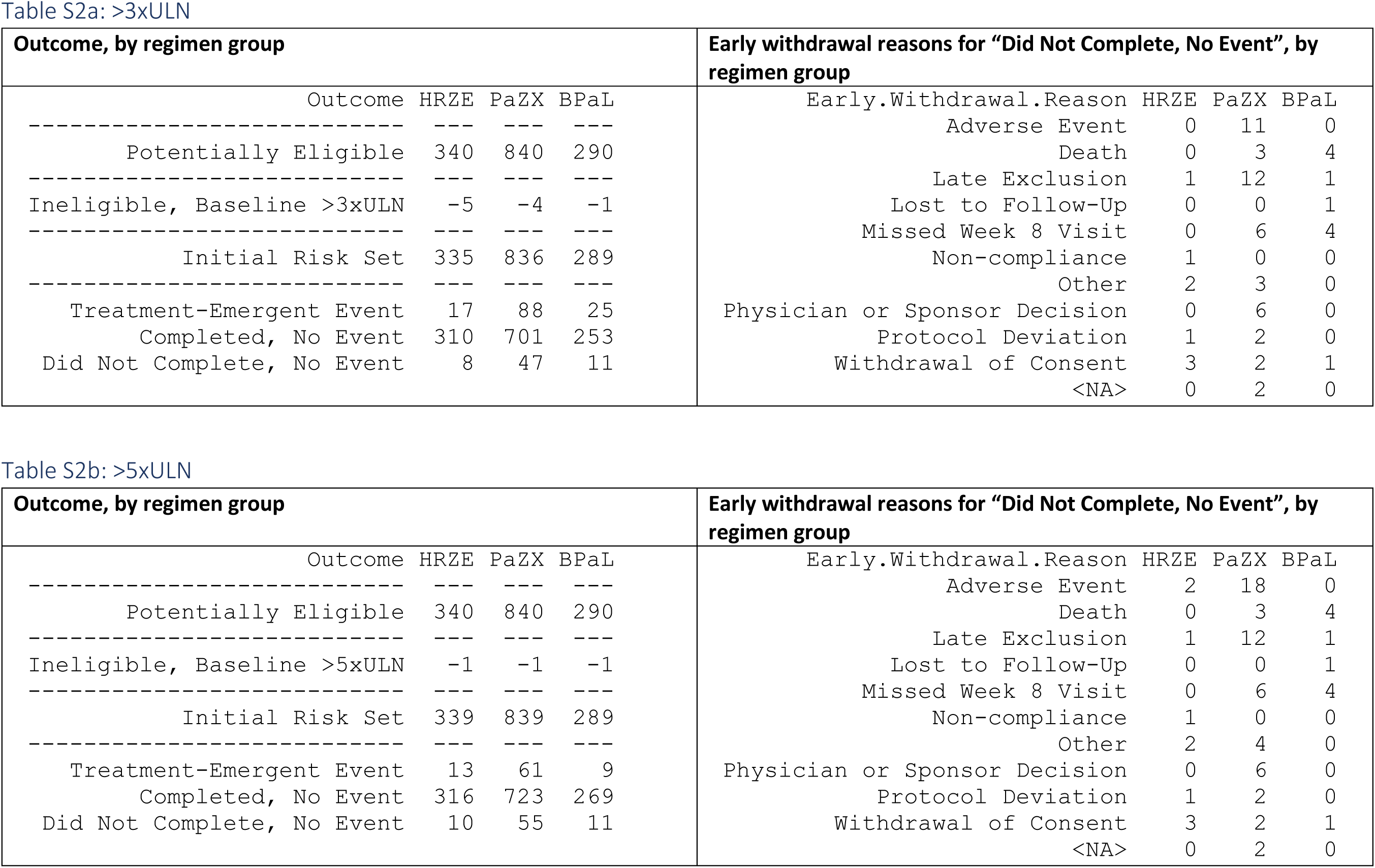
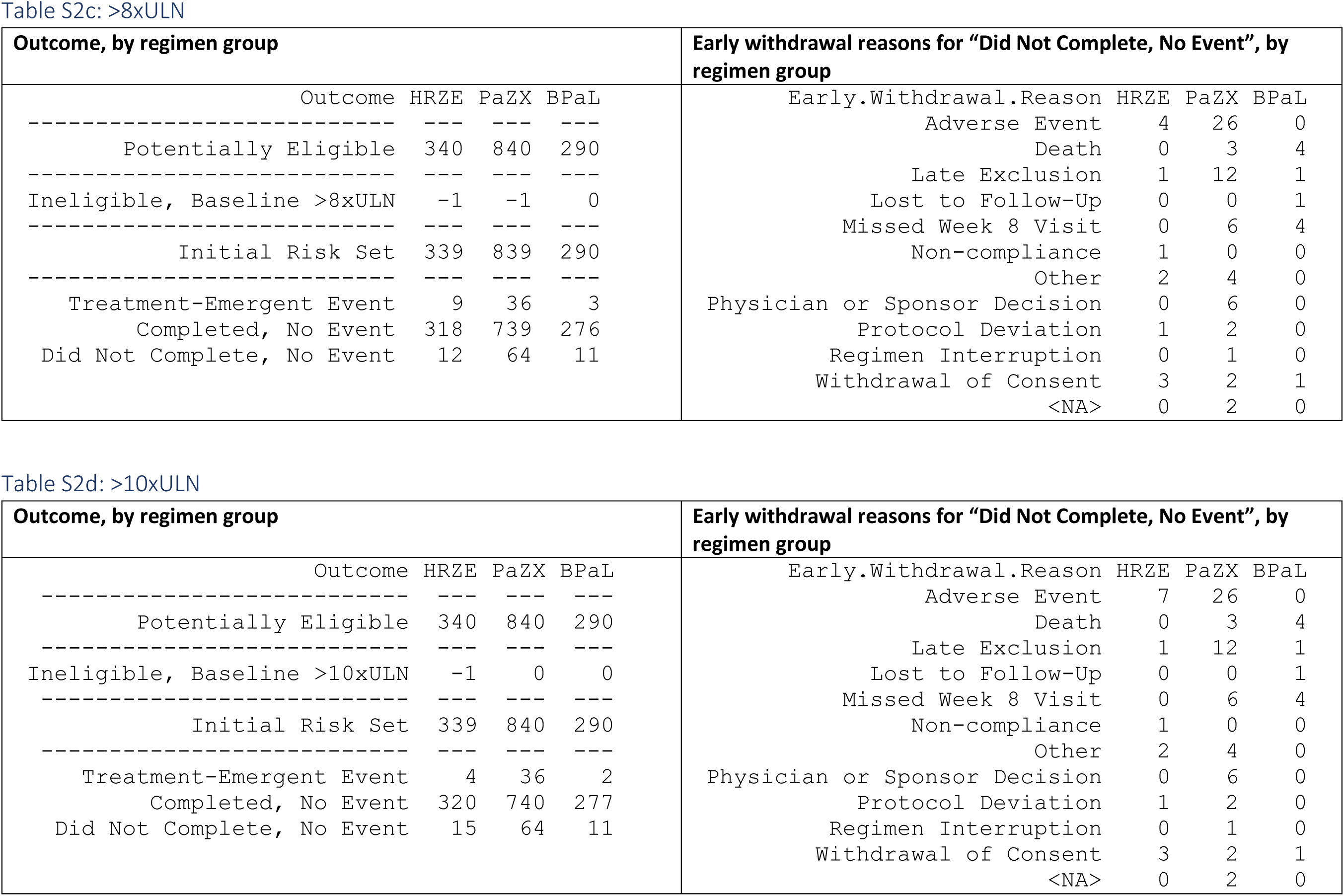
Participant Flow Through Outcomes.

**Table S3:**
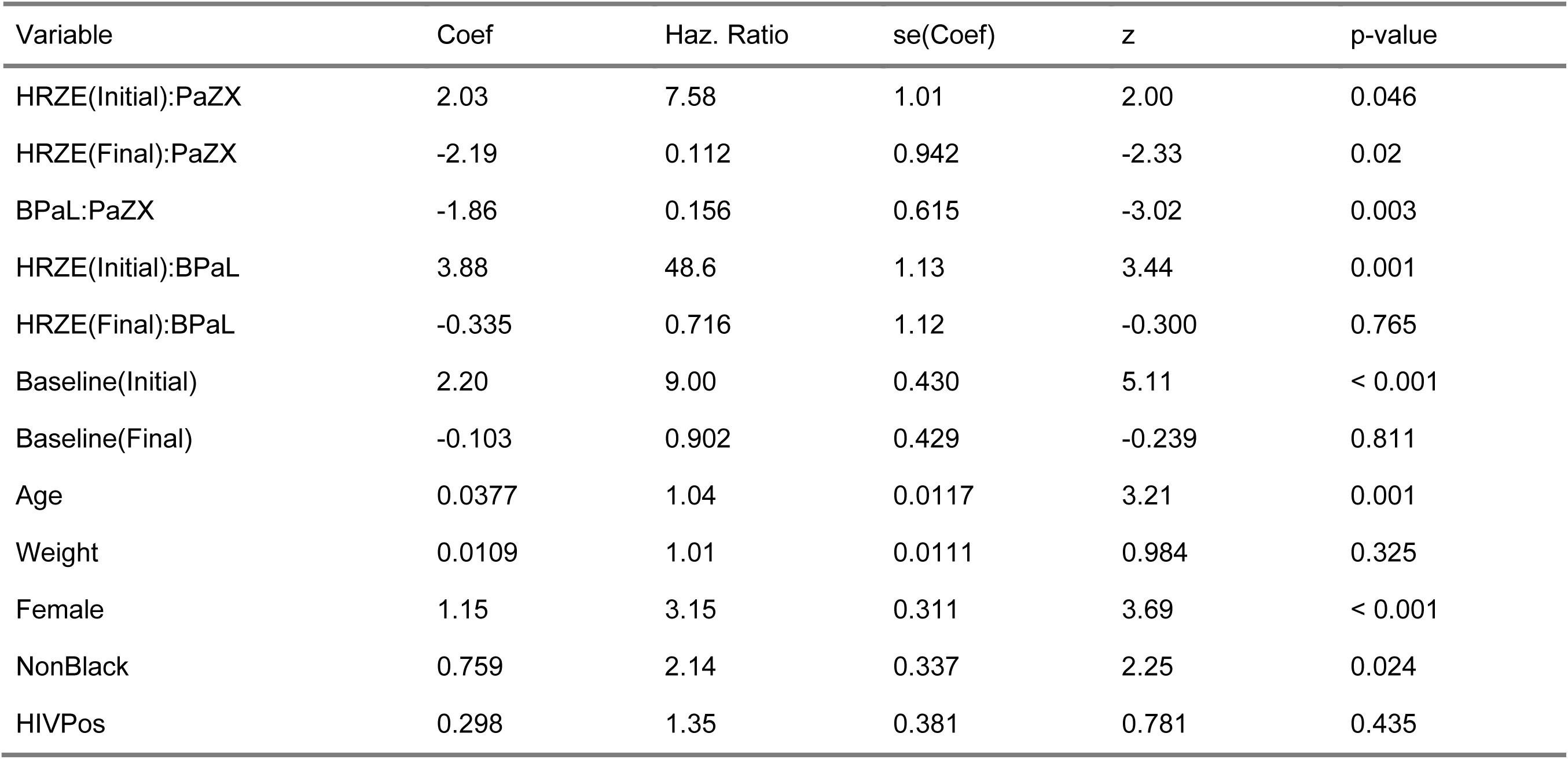
Cox Model for Time to First Elevation More than 8xULN, HRZE versus PaZX versus BPaL.

**Table S4:** Cox Model for Time to First Elevation More than 3xULN, PaZX-DS versus PaZX DR versus BPaL.

| Variable | Coef | Haz. Ratio | se(Coef) | Z | p-value |
| --- | --- | --- | --- | --- | --- |
| PaZX-DS:PaZX-DR | 0.599 | 1.82 | 0.270 | 2.22 | 0.026 |
| BPAL:PaZX-DR | -0.176 | 0.839 | 0.320 | -0.549 | 0.583 |
| PaZX-DS:BPAL | 0.775 | 2.17 | 0.255 | 3.03 | 0.002 |
| Baseline | 0.648 | 1.91 | 0.186 | 3.48 | 0.001 |
| Age | 0.0166 | 1.02 | 0.00820 | 2.02 | 0.043 |
| Weight | 0.0142 | 1.01 | 0.00731 | 1.94 | 0.052 |
| Female | 0.427 | 1.53 | 0.209 | 2.04 | 0.042 |
| NonBlack | 0.506 | 1.66 | 0.223 | 2.27 | 0.023 |
| HIVPos | 0.308 | 1.36 | 0.253 | 1.22 | 0.224 |
| AUC | 0.00137 | 1.00 | 0.00321 | 0.427 | 0.670 |

**Table S5:**
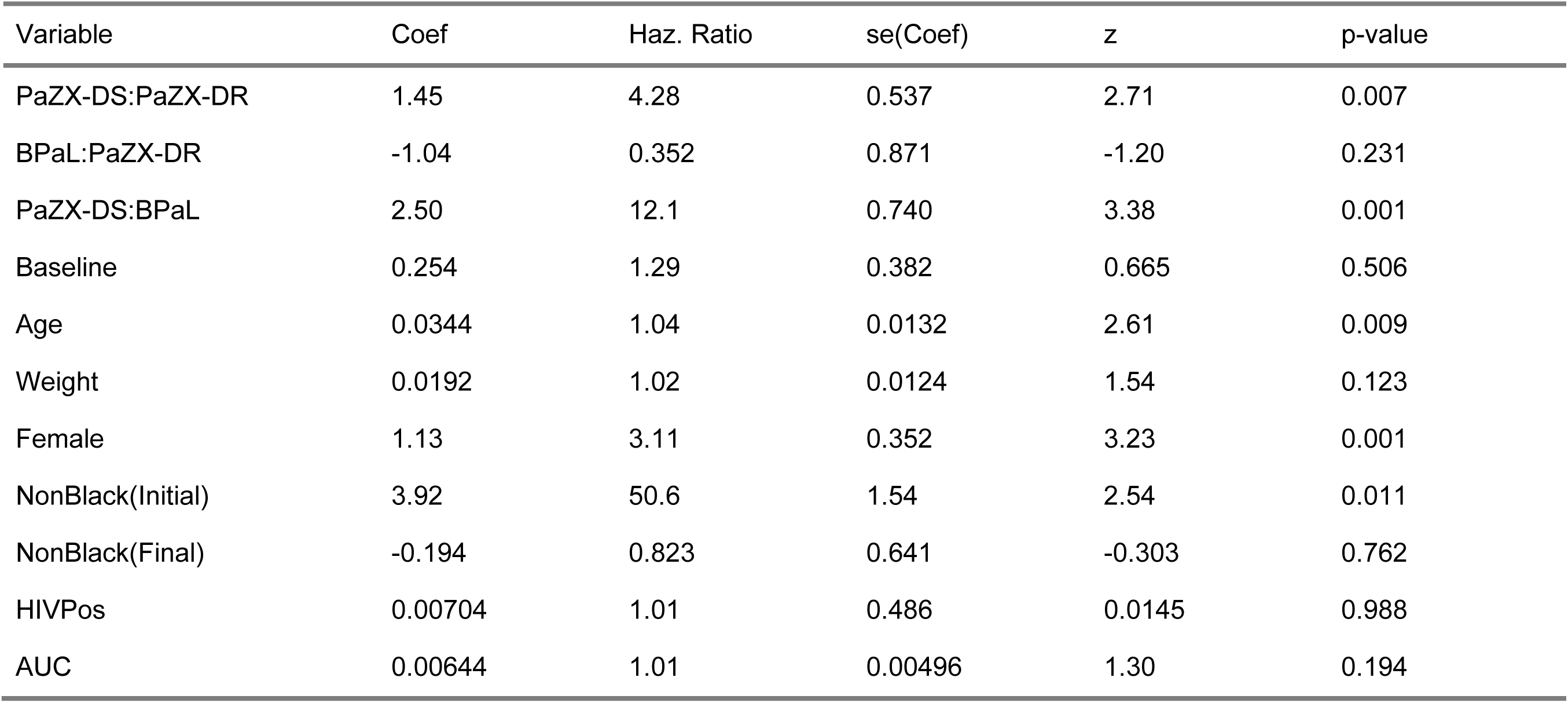
Cox Model for Time to First Elevation More than 8xULN, PaZX-DS versus PaZX-DR versus BPaL.

| Variable | Coef | Haz. Ratio | se(Coef) | z | p-value |
| --- | --- | --- | --- | --- | --- |
| PaZX-DS:PaZX-DR | 1.45 | 4.28 | 0.537 | 2.71 | 0.007 |
| BPAL:PaZX-DR | -1.04 | 0.352 | 0.871 | -1.20 | 0.231 |
| PaZX-DS:BPAL | 2.50 | 12.1 | 0.740 | 3.38 | 0.001 |
| Baseline | 0.254 | 1.29 | 0.382 | 0.665 | 0.506 |
| Age | 0.0344 | 1.04 | 0.0132 | 2.61 | 0.009 |
| Weight | 0.0192 | 1.02 | 0.0124 | 1.54 | 0.123 |
| Female | 1.13 | 3.11 | 0.352 | 3.23 | 0.001 |
| NonBlack(Initial) | 3.92 | 50.6 | 1.54 | 2.54 | 0.011 |
| NonBlack(Final) | -0.194 | 0.823 | 0.641 | -0.303 | 0.762 |
| HIVPos | 0.00704 | 1.01 | 0.486 | 0.0145 | 0.988 |
| AUC | 0.00644 | 1.01 | 0.00496 | 1.30 | 0.194 |

**Table S6:** Cox Model for Time to First Elevation More than 3xULN, HRZE versus PaZX-DS.

| Variable | Coef | Haz. Ratio | se(Coef) | z | p-value |
| --- | --- | --- | --- | --- | --- |
| HRZE:PaZX-DS | -0.855 | 0.425 | 0.271 | -3.15 | 0.002 |
| Baseline | 0.398 | 1.49 | 0.242 | 1.65 | 0.099 |
| Age | 0.0187 | 1.02 | 0.00904 | 2.07 | 0.039 |
| Weight | 0.0171 | 1.02 | 0.00794 | 2.15 | 0.031 |
| Female | 0.435 | 1.55 | 0.232 | 1.88 | 0.06 |
| NonBlack(Initial) | 2.14 | 8.50 | 0.719 | 2.98 | 0.003 |
| NonBlack(Final) | -0.0754 | 0.927 | 0.432 | -0.175 | 0.861 |
| HIVPos | 0.740 | 2.10 | 0.265 | 2.80 | 0.005 |

**Table S7:**
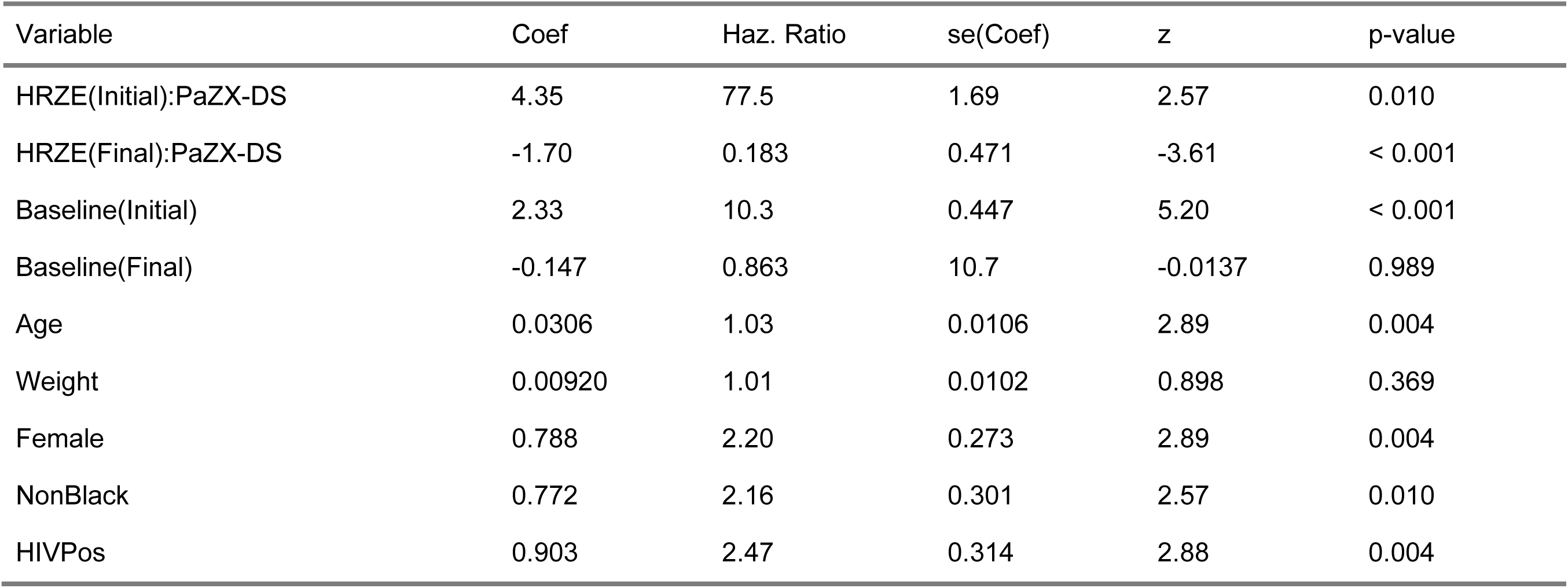
Cox Model for Time to First Elevation More than 8xULN, HRZE versus PaZX-DS.

| Variable | Coef | Haz. Ratio | se(Coef) | z | p-value |
| --- | --- | --- | --- | --- | --- |
| HRZE(Initial):PaZX-DS | 4.35 | 77.5 | 1.69 | 2.57 | 0.010 |
| HRZE(Final):PaZX-DS | -1.70 | 0.183 | 0.471 | -3.61 | < 0.001 |
| Baseline(Initial) | 2.33 | 10.3 | 0.447 | 5.20 | < 0.001 |
| Baseline(Final) | -0.147 | 0.863 | 10.7 | -0.0137 | 0.989 |
| Age | 0.0306 | 1.03 | 0.0106 | 2.89 | 0.004 |
| Weight | 0.00920 | 1.01 | 0.0102 | 0.898 | 0.369 |
| Female | 0.788 | 2.20 | 0.273 | 2.89 | 0.004 |
| NonBlack | 0.772 | 2.16 | 0.301 | 2.57 | 0.010 |
| HIVPos | 0.903 | 2.47 | 0.314 | 2.88 | 0.004 |

**Table S8:** Treatment-Emergent Potential Hy’s Law Cases.

| Study | HRZE | PaZX | BPAL |
| --- | --- | --- | --- |
| NC-002 | 1 (1.7%) |  |  |
| NC-005 | 1 (1.6%) | 1 (0.56%) |  |
| STAND | 1 (1.5%) | 5<sup>a,b</sup> (2.3%) |  |
| Nix-TB | - | - | 1<sup>c</sup> (0.92%) |
| ZeNix | - | - | 1<sup>d</sup> (0.55%) |
| SimpliciTB | 0 (0%) | 1 (0.33%) |  |
| Total | 3 (0.88%) | 7 (0.83%) | 2 (0.69%) |

<sup>a</sup>Includes one subject who was discontinued from Pa<sub>200</sub>MZ on Day 125 and started on HRZE with the NTP on Day 154; the first occurrence of BILI > 2xULN was Day 186.
<sup>b</sup>Includes one subject whose ALT and BILI elevations were reported only by local labs, not contained in study central lab database, but were hard-coded into this analysis dataset. See Narrative Case 3.
<sup>c</sup>This participant had a medical history of alcohol use. Potential-Hy's-Law lab criteria were met per central lab results at Week 8, with ALP 1.8 xULN. A local lab repeated 4 days later showed ALP 2.5 xULN. The BPAL regimen was rechallenged later, and the participant was able to complete the remaining 4 months of treatment (6-month full treatment course) with no issues.
<sup>d</sup>This participant met the potential-Hy's-Law lab criteria at study Day 183, which was attributed to documented acute hepatitis B.

## Ethical oversight of studies

### NC-002

#### Ethics statement from Clinical Study Report

The protocol, written trial patient information, informed consent forms, and any other appropriate trial-related information were reviewed and approved by an independent ethics committee (IEC) at each trial center.

#### Ethics committees and institutional review boards

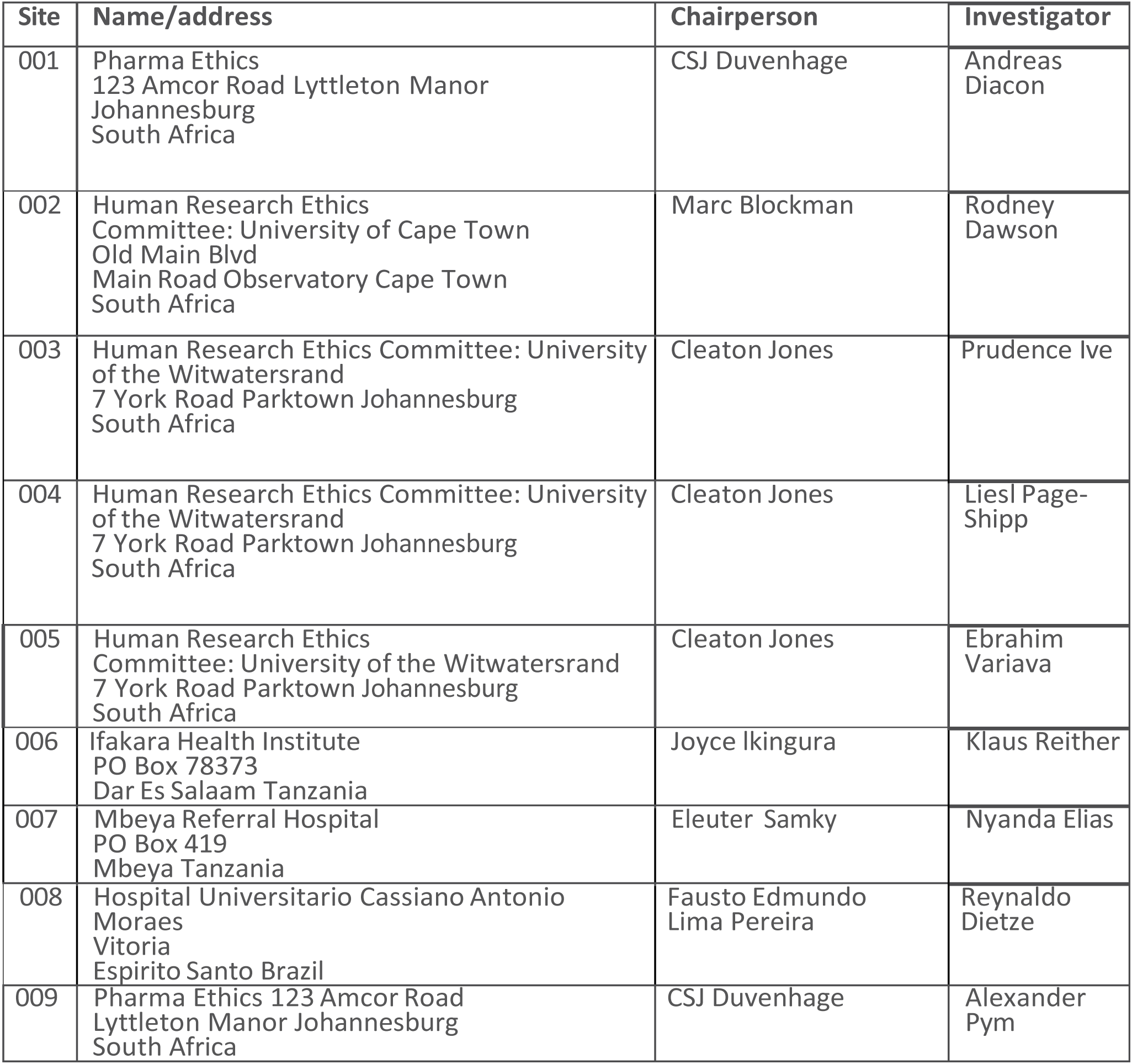

| Site | Name/address | Chairperson | Investigator |
| --- | --- | --- | --- |
| 001 | Pharma Ethics<br>123 Amcor Road Lyttleton Manor<br>Johannesburg<br>South Africa | CSJ Duvenhage | Andreas Diacon |
| 002 | Human Research Ethics<br>Committee: University of Cape Town<br>Old Main Blvd<br>Main Road Observatory Cape Town<br>South Africa | Marc Blockman | Rodney Dawson |
| 003 | Human Research Ethics Committee: University<br>of the Witwatersrand<br>7 York Road Parktown Johannesburg<br>South Africa | Cleaton Jones | Prudence Ive |
| 004 | Human Research Ethics Committee: University<br>of the Witwatersrand<br>7 York Road Parktown Johannesburg<br>South Africa | Cleaton Jones | Liesl Page-Shipp |
| 005 | Human Research Ethics<br>Committee: University of the Witwatersrand<br>7 York Road Parktown Johannesburg<br>South Africa | Cleaton Jones | Ebrahim Variava |
| 006 | Ifakara Health Institute<br>PO Box 78373<br>Dar Es Salaam Tanzania | Joyce Ikingura | Klaus Reither |
| 007 | Mbeya Referral Hospital<br>PO Box 419<br>Mbeya Tanzania | Eleuter Samky | Nyanda Elias |
| 008 | Hospital Universitario Cassiano Antonio<br>Moraes<br>Vitoria<br>Espirito Santo Brazil | Fausto Edmundo<br>Lima Pereira | Reynaldo Dietze |
| 009 | Pharma Ethics 123 Amcor Road<br>Lyttleton Manor Johannesburg<br>South Africa | CSJ Duvenhage | Alexander Pym |

### NC-003

#### Ethics statement from Clinical Study Report

The protocol, written trial patient information, informed consent forms, and any other appropriate trial-related information were reviewed and approved by an independent ethics committee (IEC) at each trial center.

#### Ethics committees and institutional review boards

| Site | Name/address | Chairperson | Investigator |
| --- | --- | --- | --- |
| 001 | Pharma Ethics<br>123 Amcor Road Lyttleton Manor<br>Johannesburg<br>South Africa | CSJ<br>Duvenhage | Andreas<br>Diacon |
| 002 | Human Research Ethics<br>Committee: University of Cape Town<br>Old Main Blvd<br>Main Road Observatory Cape Town<br>South Africa | Marc<br>Blockman | Rodney<br>Dawson |

### NC-005

#### Ethics statement from Clinical Study Report

The protocol, protocol amendment, written trial patient information and informed consent forms (ICFs), and any other appropriate trial-related information including the Investigator’s Brochures were reviewed and approved by an independent ethics committee (IEC) in South Africa and Uganda, and an institutional review board (IRB) in Tanzania.

#### Ethics committees and institutional review boards

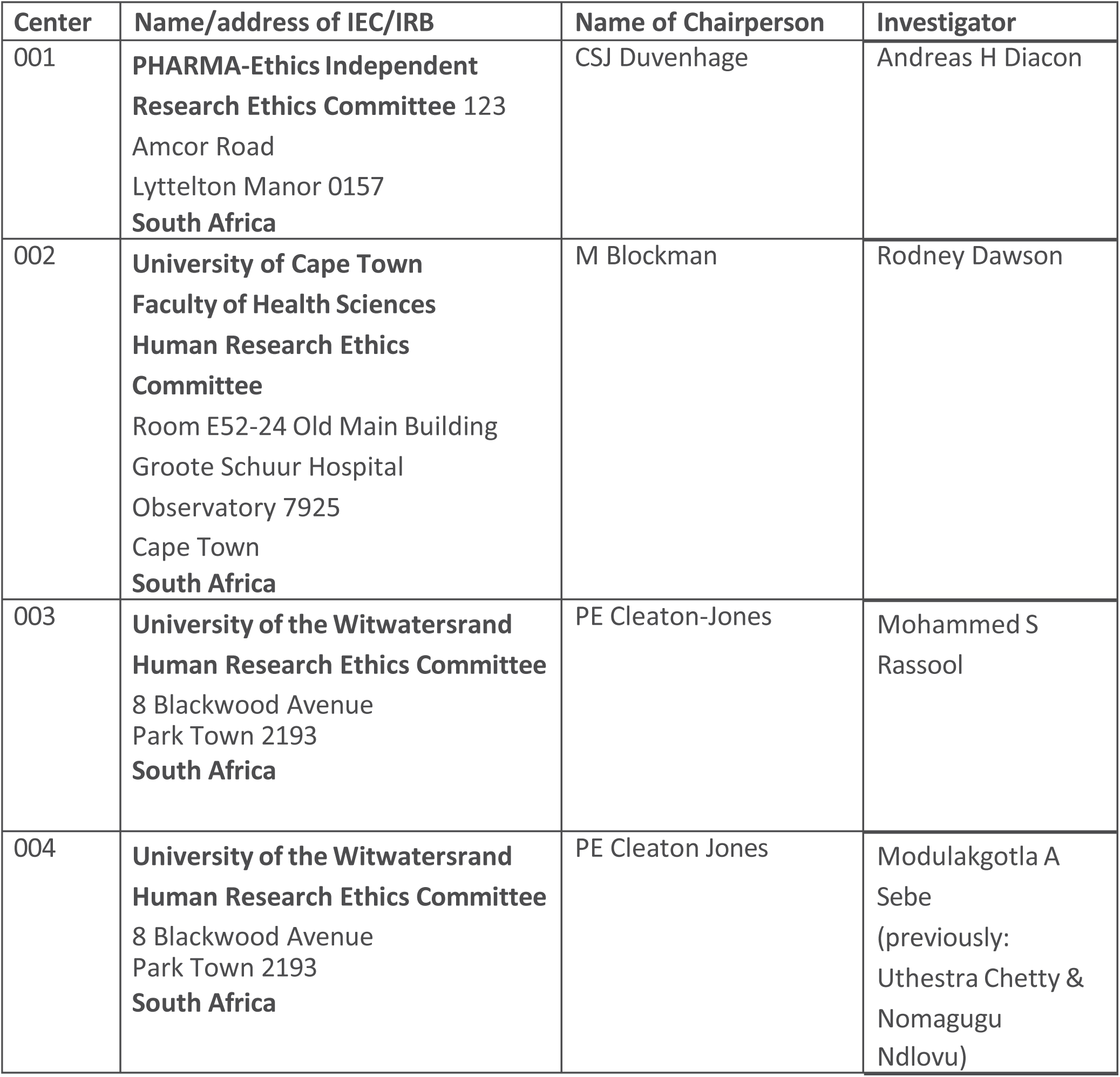
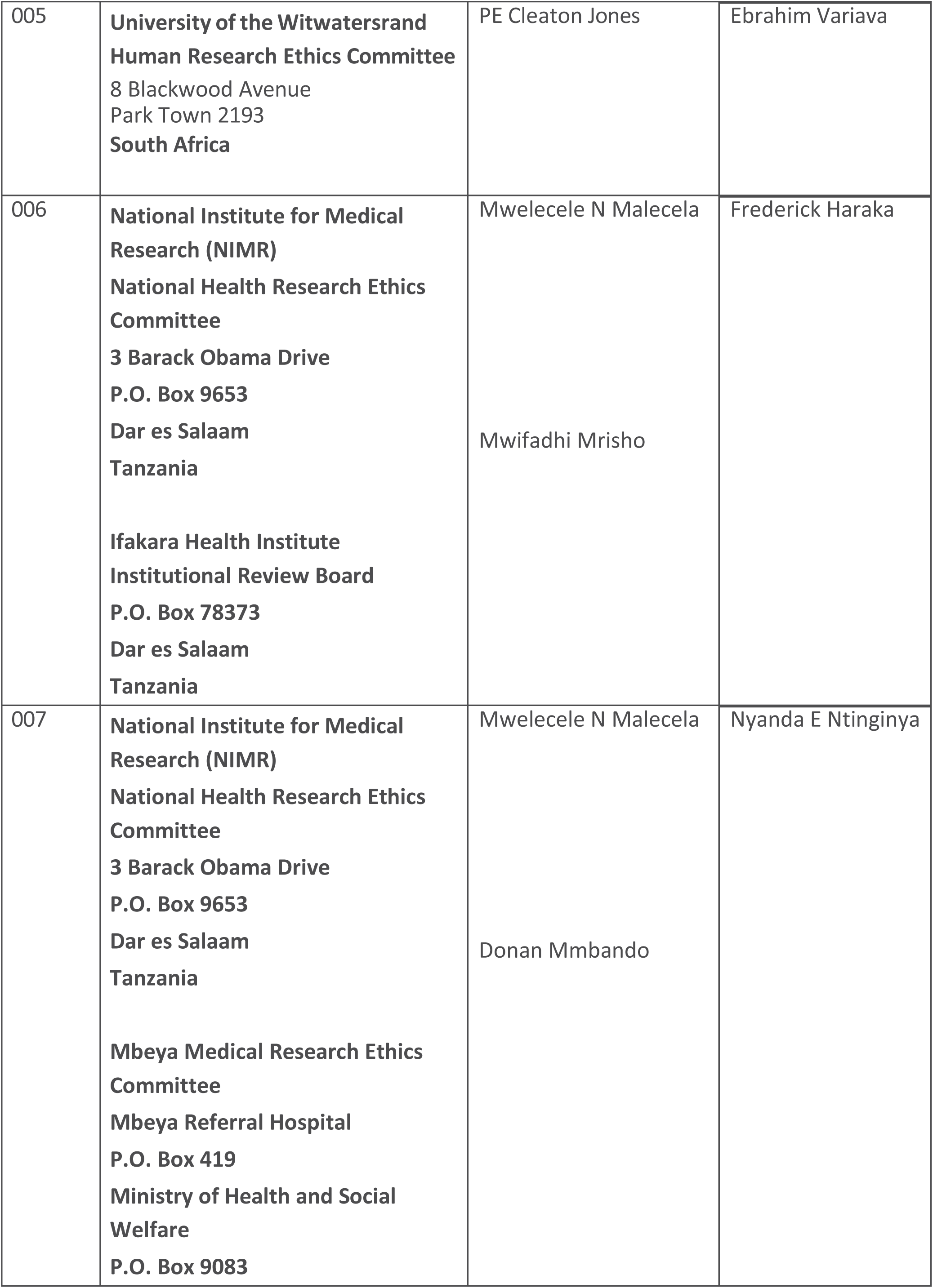
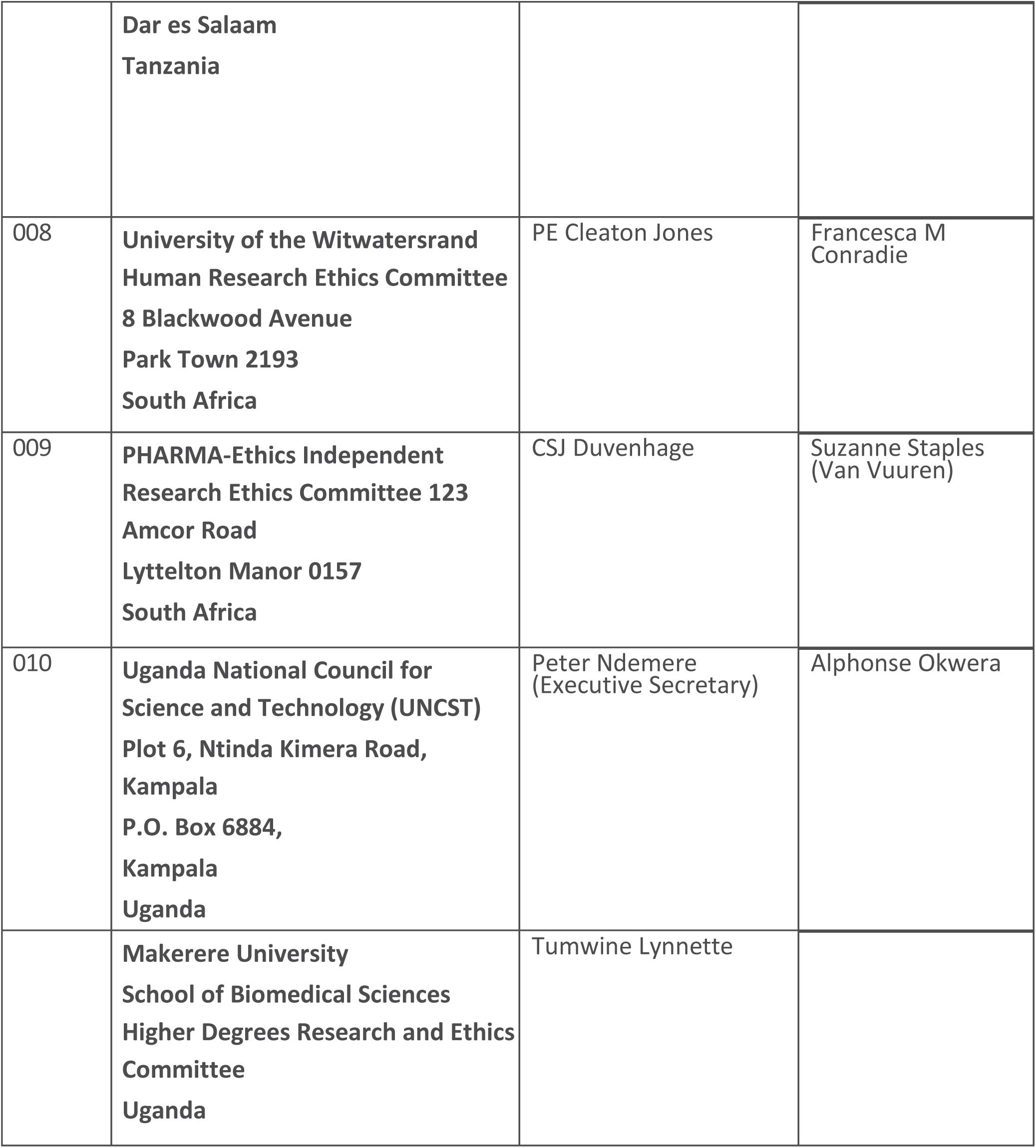

### STAND

#### Ethics statement from Clinical Study Report

The trial protocol, all trial protocol amendments, written trial patient information, informed consent form (ICF), Investigator’s Brochure (IB) and any other relevant documents were reviewed and approved by an Independent Ethics Committee (IEC)/Institutional Review Board (IRB) at each trial center.

#### Ethics committees and institutional review boards

| **Trial Center** | **Name / Address** |
| --- | --- |
| 0201 | Medical Research Ethics Committee<br>Ministry of Health Malaysia<br>c/o National Institute of Health Secretariat, Institute for Health Management, Jalan Rumah Sakit, Bangsar<br>59000 Kuala Lumpur, Malaysia, |
| 0203 | UKM Research and Ethics Committee<br>Bangunan Wawasan<br>Universiti Kebangsaan Malaysia<br>Selangor, 43600, Malaysia |
| 0204 | Research Management Institute (RMI)<br>Universiti Teknologi MARA (UiTM)<br>Selayang Campus, Jalan Prima Selayang 7<br>Shah Alam, Selangor, 68100, Malaysia |
| 0301 | University of Cape Town Human Research Ethics Committee<br>Room E52-24 Old Main Building, Groote Schuur Hospital, Observatory<br>Cape Town, 7925, Cape Province, South Africa |
| 0302<br>0303<br>0304<br>0305<br>0307<br>0308<br>0309<br>0310<br>0311<br>0312<br>0313 | University of Witwatersrand Human Research Ethics Committee<br>(Medical)<br>Suite 189, Private Bag x2600, Houghton<br>Johannesburg, 2041, Gauteng, South Africa |
| 0401 | Mbeya Medical Research and Ethics Committee<br>Mbeya Consultant Hospital<br>PO Box 419, Mbeya, Tanzania<br>National Institute of Medical Research<br>3 Barack Obama Drive, PO Box 9653, 11101<br>Dar-es-Salaam, Tanzania |
| 0402 | Ifakara Health Institute Institutional Review Board<br>PO Box 78373, Dar-es-Salaam, Tanzania<br>National Institute of Medical Research<br>3 Barack Obama Drive, PO Box 9653, 11101<br>Dar-es-Salaam, Tanzania |
| 0501 | Central Research Ethics Committee (CREC)<br>Central Ethics Committee<br>National Research Council of Thailand<br>196 Phahonyothin Road, Lat Yao, Chatuchak<br>Bangkok, 10900, Thailand<br>Ethical Review Committee of Central Chest Institute of Thailand<br>Local Ethics Committee<br>Ninth Building, Fifth Floor<br>Tiwanon Road, Bangkasor Sub-district<br>Muang, Nonthaburi, 11000, Thailand<br>Institute Review Board of Bamrasnaradura Institute<br>Infectious Diseases Institute<br>Local Ethics Committee<br>126 Tiwanont Road<br>Muang, Nonthaburi, 11000, Thailand |
| 0601 | LEC Mun. Tuberculosis Hospital #1<br>Local Ethics Committee<br>121/3, Kharkivske Shose Street<br>Kyiv, 02091, Ukraine |
| 0602 | LEC SI National Institute of Phthisiology and Pulmonology<br>named F.G.Yanovskyy of AMS of Ukraine<br>Local Ethics Committee<br>3, F. Rakotcy Street<br>Uzhgorod, 88000, Ukraine |
| 0701 | Kenya Medical Research Institute Scientific Ethics Review Unit<br>(KEMRI SERU)<br>PO Box 54840-00200<br>Nairobi, Kenya |
| 0702 | Kenya Medical Research Institute Scientific Ethics Review Unit (KEMRI SERU)<br>PO Box 54840-00200<br>Nairobi, Kenya |
| 1002 | University of the Philippines Manila Research Ethics Board<br>Local Ethics Committee<br>Second Floor Paz Mendoza Building<br>College of Medicine UP<br>547 Pedro Gil Street, Ermita, Manila, 1000, Philippines |
| 1003 | Lung Center of The Philippines<br>Local Ethics - Ethics Committee<br>Lung Center of The Philippines<br>Quezon Avenue, Diliman<br>Quezon City, 1100, Philippines<br>Lung Center of the Philippines<br>Local Ethics - Technical Review Board<br>Lung Center of The Philippines<br>Quezon Avenue, Diliman<br>Quezon City, 1100, Philippines |
| 1004 | Tropical Disease Foundation Institutional Review Board<br>Local Ethics Committee<br>Amorsolo Street Corner Urban Avenue, Pio del Pilar<br>Philippine Institute of Tuberculosis Building<br>Makati City, 1230, Philippines |
| 1301 | University of Makerere Scientific and Biomedical Studies Institutional Review Board<br>PO Box 7072<br>Kampala, Uganda<br>Uganda National Council for Science and Technology (UNCST)<br>Plot 6 Kimera Road, PO Box 6884<br>Nitinda |
| 1501 | LEC of National Center for Tuberculosis and Lung Diseases<br>Local Ethics<br>50, Maruashvili Street<br>Tbilisi, 0101, Georgia |

### Nix-TB

#### Ethics statement from Clinical Study Report

The trial protocol, all trial protocol amendments, written trial patient information, Informed Consent Form (ICF), Investigator’s Brochure (IB), and any other relevant documents were reviewed and approved by an Independent Ethics Committee (IEC) or Institutional Review Board (IRB) at each trial center.

#### Ethics committees and institutional review boards

| **Trial Center** | **IEC Name / Address** | **IEC Chairperson** |
| --- | --- | --- |
| 001 | The University of the Witwatersrand<br>Human Research Ethics Committee:<br>(Medical) / 8 Blackwood Avenue,<br>Parktown, Johannesburg, 2193, Gauteng,<br>South Africa, , Fax: | Professor PE Cleaton-Jones |
| 002 | Pharma-Ethics (Pty) Ltd / 123 Amcor<br>Road, Lyttleton Manor, Pretoria, 0157,<br>Gauteng, South Africa,<br>8690, | Dr CSJ Duvenhage |
| 004 | The University of the Witwatersrand<br>Human Research Ethics Committee:<br>(Medical) / 8 Blackwood Avenue,<br>Parktown, Johannesburg, 2193, Gauteng,<br>South Africa, , Fax: | Professor PE Cleaton-Jones |

### ZeNix

#### Ethics statement from Clinical Study Report

The protocol and required trial-related documents were reviewed by each site’s respective Independent Ethics Committee (IEC)/Institutional Review Board (IRB) and operated in accordance with International Council on Harmonisation (ICH) Good Clinical Practice (GCP), the ethical principles that have their origin in the Declaration of Helsinki. The investigator maintained an accurate and complete record of all submissions made to, and responses from, the IRB/IEC.

The trial was conducted in accordance with the protocol, the ethical principles that have their origin in the Declaration of Helsinki, and the applicable regulatory requirements.

#### Ethics committees and institutional review boards

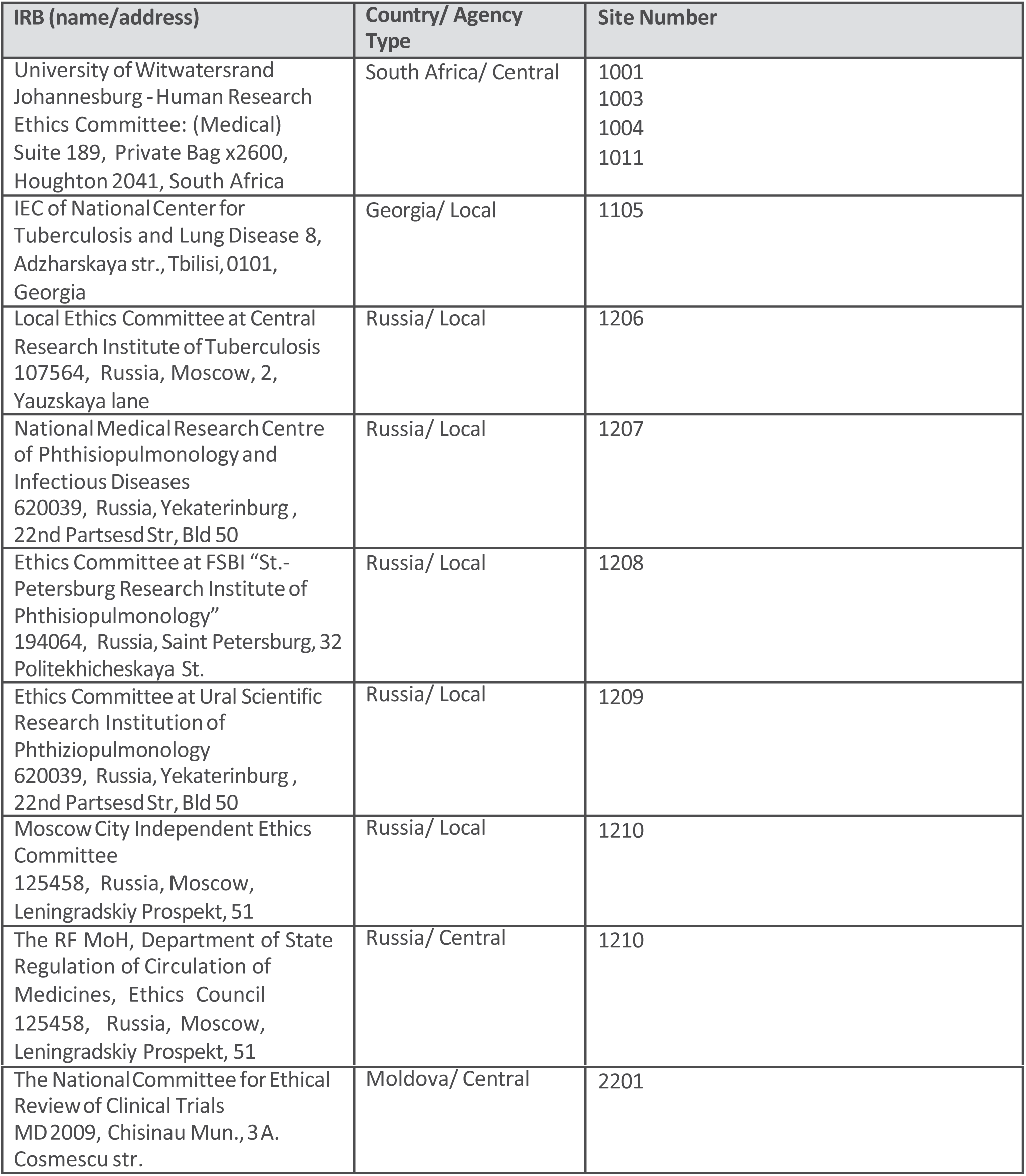

| IRB (name/address) | Country/ Agency Type | Site Number |
| --- | --- | --- |
| University of Witwatersrand<br>Johannesburg - Human Research<br>Ethics Committee: (Medical)<br>Suite 189, Private Bag x2600,<br>Houghton 2041, South Africa | South Africa/ Central | 1001<br>1003<br>1004<br>1011 |
| IEC of National Center for<br>Tuberculosis and Lung Disease 8,<br>Adzharskaya str., Tbilisi, 0101,<br>Georgia | Georgia/ Local | 1105 |
| Local Ethics Committee at Central<br>Research Institute of Tuberculosis<br>107564, Russia, Moscow, 2,<br>Yauzskaya lane | Russia/ Local | 1206 |
| National Medical Research Centre<br>of Phthisiopulmonology and<br>Infectious Diseases<br>620039, Russia, Yekaterinburg ,<br>22nd Partsesd Str, Bld 50 | Russia/ Local | 1207 |
| Ethics Committee at FSBI "St.-<br>Petersburg Research Institute of<br>Phthisiopulmonology"<br>194064, Russia, Saint Petersburg, 32<br>Politekhicheskaya St. | Russia/ Local | 1208 |
| Ethics Committee at Ural Scientific<br>Research Institution of<br>Phthisiopulmonology<br>620039, Russia, Yekaterinburg ,<br>22nd Partsesd Str, Bld 50 | Russia/ Local | 1209 |
| Moscow City Independent Ethics<br>Committee<br>125458, Russia, Moscow,<br>Leningradskiy Prospekt, 51 | Russia/ Local | 1210 |
| The RF MoH, Department of State<br>Regulation of Circulation of<br>Medicines, Ethics Council<br>125458, Russia, Moscow,<br>Leningradskiy Prospekt, 51 | Russia/ Central | 1210 |
| The National Committee for Ethical<br>Review of Clinical Trials<br>MD2009, Chisinau Mun., 3 A.<br>Cosmescu str. | Moldova/ Central | 2201 |

### SimpliciTB

#### Ethics statement from Clinical Study Report

The protocol and required trial-related documents were reviewed by each site’s respective Independent Ethics Committee (IEC)/Institutional Review Board (IRB). The trial did not start until the IEC/IRB had approved the protocol, written informed consent, any written information to be provided to the participant or any modification thereof, plus any other trial-related documents required for review. The IEC/IRB was constituted and operated in accordance with International Council on Harmonisation (ICH) Good Clinical Practice (GCP) and the ethical principles that have their origin in the Declaration of Helsinki. The investigator maintained an accurate and complete record of all submissions made to, and responses from, the IEC/IRB. The records were filed in the investigator’s trial file, and copies were sent to the sponsor.

#### Ethics committees and institutional review boards

| IRB (Name/address) | Country/Agency Type | Site Number |
| --- | --- | --- |
| University of Witwatersrand<br>Johannesburg - Human Research<br>Ethics Committee: (Medical)<br>Cnr York Road and 29 Princess of<br>Wales Terrace<br>Phillip Tobias Building Offices<br>301-304, 3rd Floor<br>Parktown, Gauteng 2193, South<br>Africa | South Africa/Central | 1001<br>1002<br>1003<br>1005<br>1006<br>1007<br>1008<br>1009<br>1010<br>1011 |
| University of Cape Town, Faculty of<br>Health Sciences Human Research<br>Ethics Committee<br>Main Road, Groote Schuur Hospital<br>Room E52.23<br>Old Main Building<br>Cape Town, Western Cape 7925,<br>South Africa | South Africa/Local | 1004 |
| IEC of National Center for<br>Tuberculosis and Lung Disease<br>Maruashvili Street 50<br>Tbilisi 0101, Georgia | Georgia/Local | 1105 |
| The RF MoH, Department of State<br>Regulation of Circulation of<br>Medicines, Ethics Council<br>GSP 4, Rakhmanovskiy Pereulok 3<br>Tverskoy, Moscow 127051, Russian<br>Federation | Russia/Central | 1206<br>1207<br>1208<br>1209<br>1210 |
| Local Ethics Committee at Central<br>Research Institute of Tuberculosis<br>2, Yauzskaya lane<br>Moscow 107564, Russian Federation | Russia/Local | 1206 |
| Ethics Committee at Ural Scientific Research Institution of Phthiziopulmonology<br>Ulitsa 22 Partsiezda, 50<br>Yekaterinburg, 620039, Russian Federation | Russia/Local | 1207 |
| Ethics Committee at Ural State Medical University<br>Ulitsa Repina, 3<br>Yekaterinburg, 620019, Russian Federation | Russia/Local | 1207 |
| Ethics Committee at FSBI "St.-Petersburg Research Institute of Phthiziopulmonology" Ligovskiy av. 2-4<br>St. Petersburg 191036, Russian Federation | Russia/Local | 1208 |
| Ethics Committee at Ural Scientific Research Institution of Phthiziopulmonology<br>Ulitsa 22 Partsiezda, 50<br>Yekaterinburg 620039, Russian Federation | Russia/Local | 1209 |
| Moscow City Independent Ethics Committee<br>ulitsa Minskaya, 12, korp. 2<br>Moscow 121096, Russian Federation | Russia/Local | 1210 |
| National Health Research Ethics Review Committee<br>3 Barack Obama Drive Dar Es Salaam 11001<br>United Republic of Tanzania | Tanzania/Central | 1401<br>1402<br>1403<br>1404 |
| Kilimanjaro Christian Medical College Research Ethics and Review Committee (CRERC)<br>Longuo B, Sokoine Road<br>Kilimanjaro Christian Medical College<br>Moshi, Kilimanjaro<br>United Republic of Tanzania | Tanzania/Local | 1404 |
| Ifakara Health Institute<br>Institutional Review Board<br>Kiko Avenue<br>Ifakara Health Institute<br>Mikocheni, Dar es Salaam<br>United Republic of Tanzania | Tanzania/Local | 1402 |
| Mbeya Medical Research and Ethics Committee<br>Hospital Hill Road<br>Dar es Salaam, Mbeya 11000<br>United Republic of Tanzania | Tanzania/Local | 1401 |
| Uganda National Council for Science and Technology (UNCST)<br>Kimera Road Ntinda, Plot 6<br>Kampala, Uganda | Uganda/ Central | 1501 |
| Makerere University College of Health Sciences, School of Biomedical Sciences<br>Mulago, PO Box 7072<br>Makerere Univ. Kampala<br>College of Health Sciences, Infectious Diseases Institute (Medicine)<br>Kampala, Uganda | Uganda/Local | 1501 |
| Medical Research Ethic Committee Institute for Health Management Jalan Rumah Sakit Bangsar<br>Kuala Lumpur 50900, Malaysia | Malaysia/Central | 1601 |
| Makati Medical Center Institutional Review Board<br>2 Amorsolo Street, Legaspi Village Makati City 1229, Philippines | Philippines/Local | 1701 |
| Ethics Review Committee of the Lung Center of Philippines North Avenue<br>Quezon City 1100, Philippines | Philippines/Local | 1702 |
| Institutional Review Board - Jose R. Reyes Memorial Medical Center Rizal Avenue, Station Cruz<br>Manila 1003, Philippines | Philippines/Local | 1703 |
| Comissao Nacional de Etica em Pesquisa – CONEP<br>SRTV 701, Via W 5 Norte, Lote D – Edifício PO 700, 3º andar<br>Asa Norte<br>Brasilia, Distrito Federal 70719-040, Brazil | Brazil/Central | 2001<br>2002 |
| Comitê de Ética em Pesquisa do Instituto Nacional de Infectologia Evandro Chagas - INI – Fiocruz<br>Avenida Brasil, 4365 Prédio da Direção, 1º Andar Manguinhos<br>Rio de Janeiro 21040-900, Brazil | Brazil/Local | 2001 |
| Comitê de Ética em Pesquisa da<br>Escola Nacional de Saúde<br>Pública Sérgio Arouca – ENSP<br>Rua Leopoldo Bulhões, 1480<br>Térreo<br>Manguinhos<br>Rio de Janeiro 21041-210,<br>Brazil | Brazil/Local | 2002 |

## DMID toxicity scale 2007 draft

*Source: U.S. National Institute of Allergy and Infectious Diseases, DMID, November 2007 (Draft)*

### ABBREVIATIONS

Abbreviations utilized in the Table:

|  |  |
| --- | --- |
| ULN = Upper Limit of Normal | LLN = Lower Limit of Normal |
| R<sub>x</sub> = Therapy | Req = Required |
| Mod = Moderate | IV = Intravenous |
| ADL = Activities of Daily Living | Dec = Decreased |

### ESTIMATING SEVERITY GRADE

For abnormalities NOT found elsewhere in the Toxicity Tables use the scale below to estimate grade of severity:

**ESTIMATING SEVERITY GRADE** For abnormalities NOT found elsewhere in the Toxicity Tables use the scale below to estimate grade of severity:
| Grade | Severity Rating | Definition |
| --- | --- | --- |
| **GRADE 1** | Mild | Transient or mild discomfort (< 48 hours); no medical intervention/therapy required. |
| **GRADE 2** | Moderate | Mild to moderate limitation in activity - some assistance may be needed; no or minimal medical intervention/therapy required. |
| **GRADE 3** | Severe | Marked limitation in activity, some assistance usually required; medical intervention/therapy required, hospitalizations possible. |
| **GRADE 4** | Potentially Life-threatening | Extreme limitation in activity, significant assistance required; significant medical intervention/therapy required, hospitalization or hospice care probable. |

### SERIOUS OR LIFE-THREATENING AEs

ANY clinical event deemed by the clinician to be serious or life-threatening should be considered a grade 4 event. Clinical events considered to be serious or life-threatening include, but are not limited to: seizures, coma, tetany, diabetic ketoacidosis, disseminated intravascular coagulation, diffuse petechiae, paralysis, acute psychosis, severe depression.

#### COMMENTS REGARDING THE USE OF THESE TABLES

- Standardized and commonly used toxicity tables (Division of AIDS, NCI’s Common Toxicity Criteria (CTC), and World Health Organization (WHO)) have been adapted for use by the Division of Microbiology and Infectious Diseases (DMID) and modified to better meet the needs of subjects in DMID trials.
- For parameters not included in the following Toxicity Tables, sites should refer to the “Guide

For Estimating Severity Grade” located above.

- Criteria are generally grouped by body system.
- Some protocols may have additional protocol specific grading criteria, which will supersede the use of these tables for specified criteria.

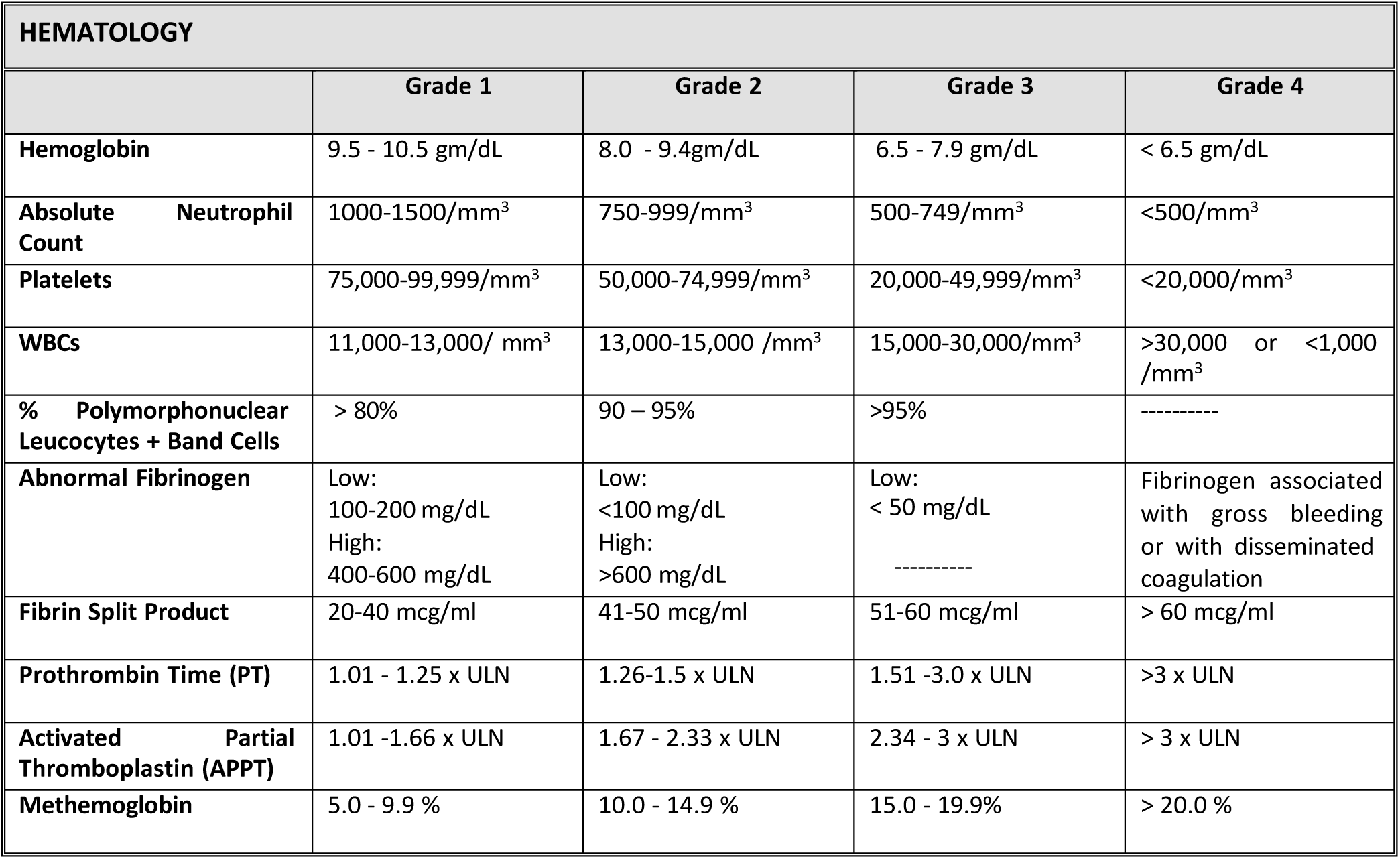
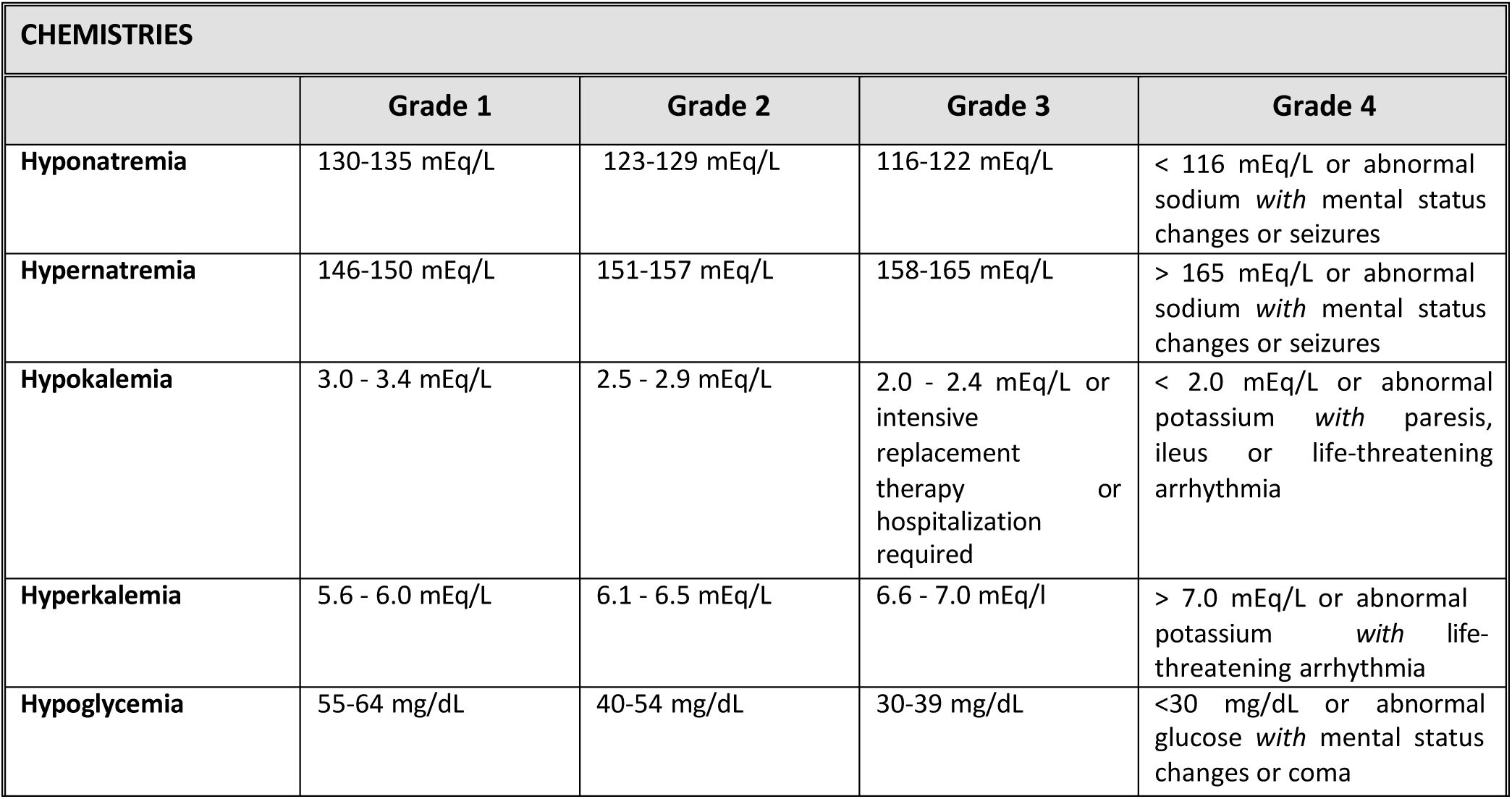
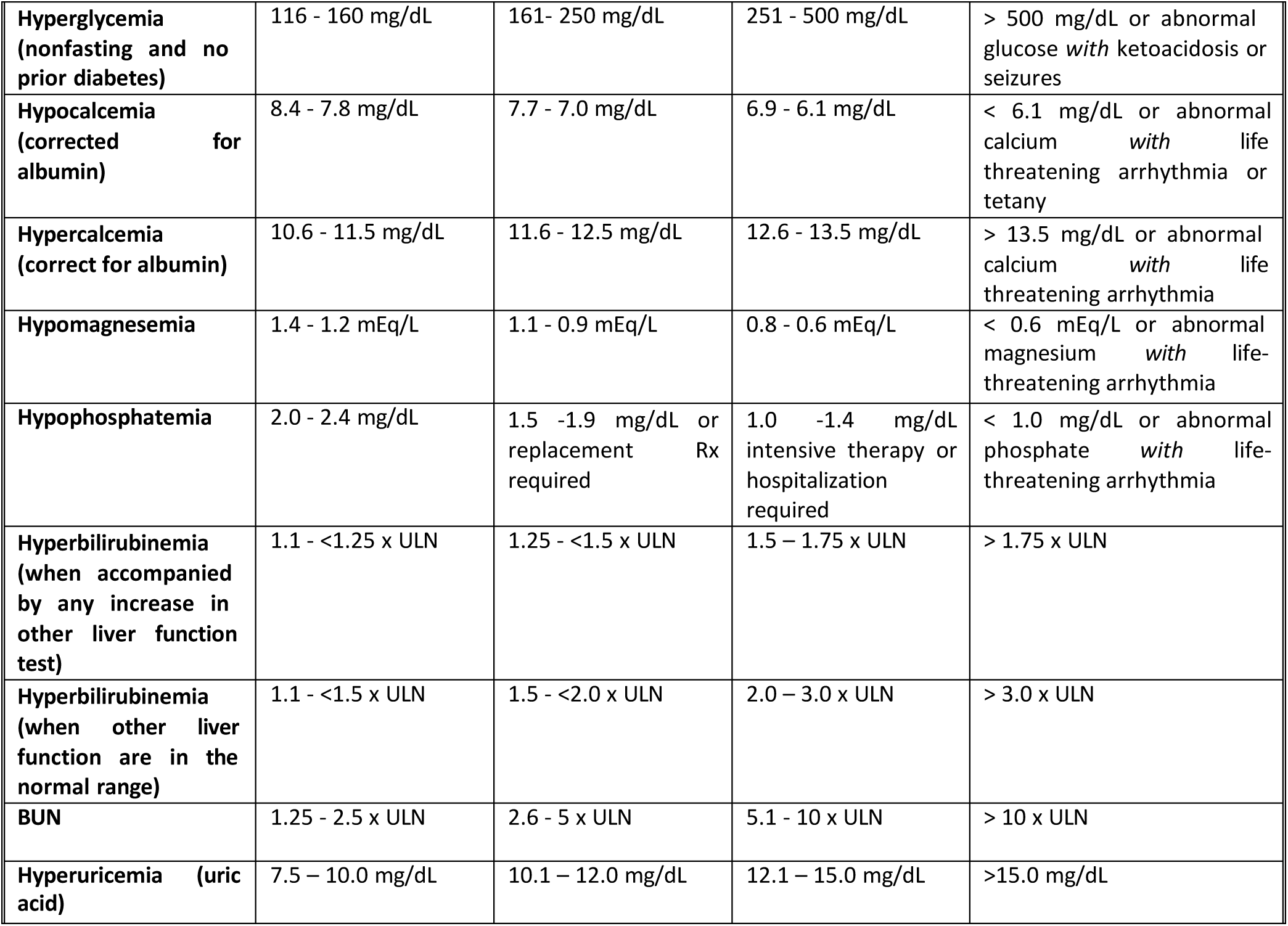
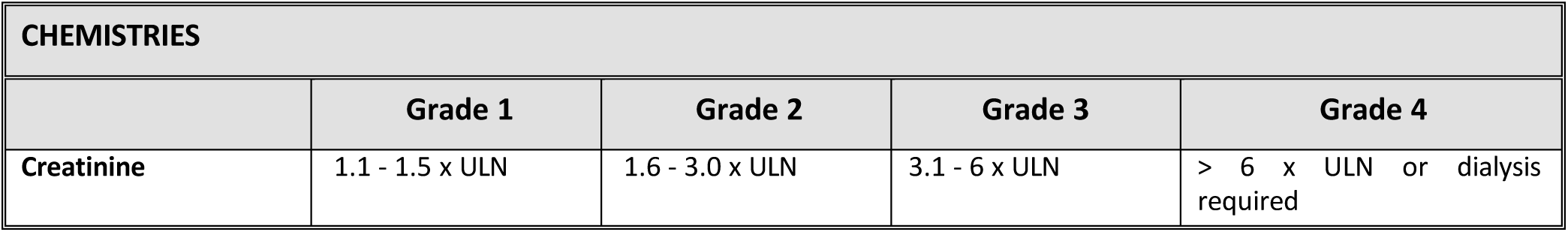
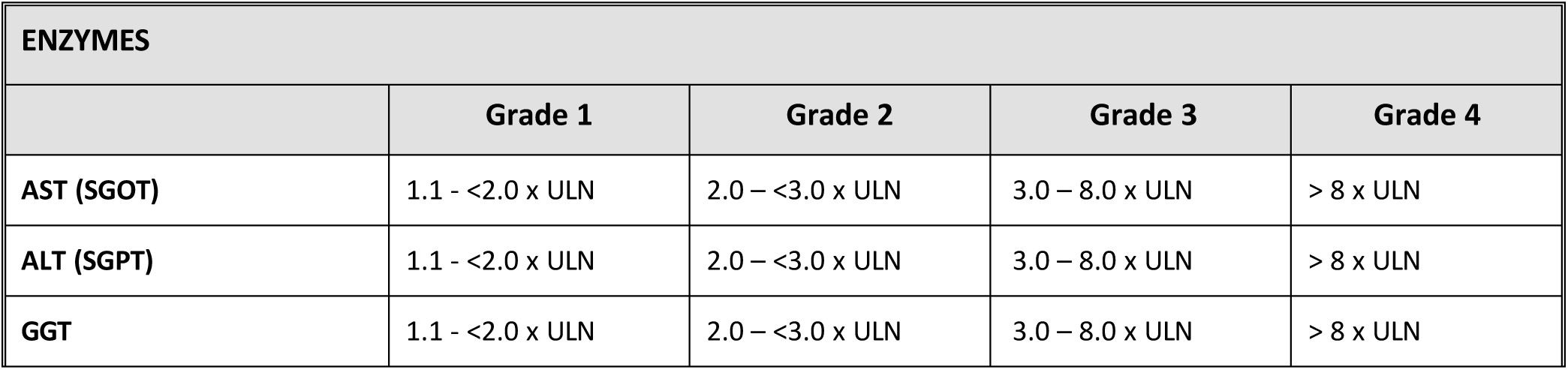
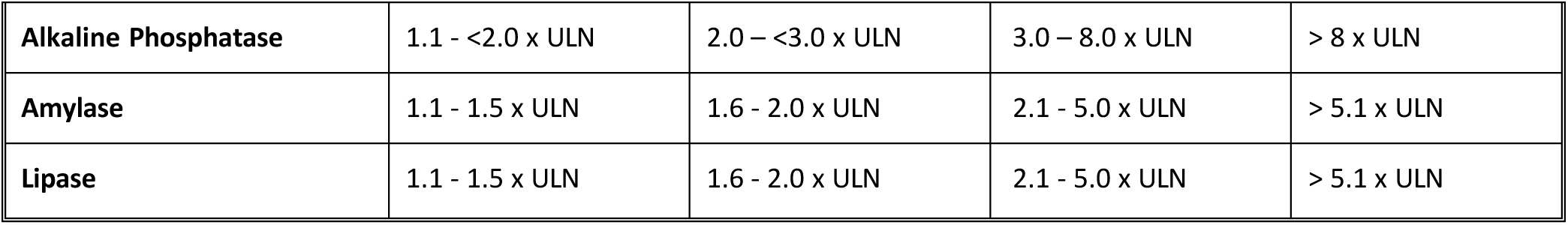
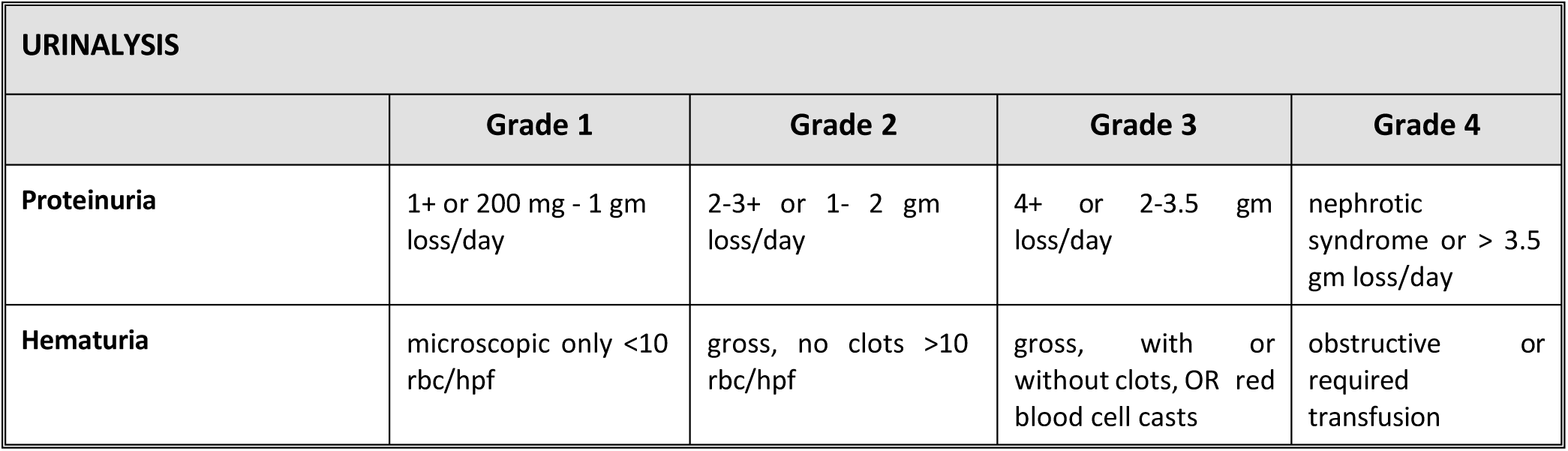
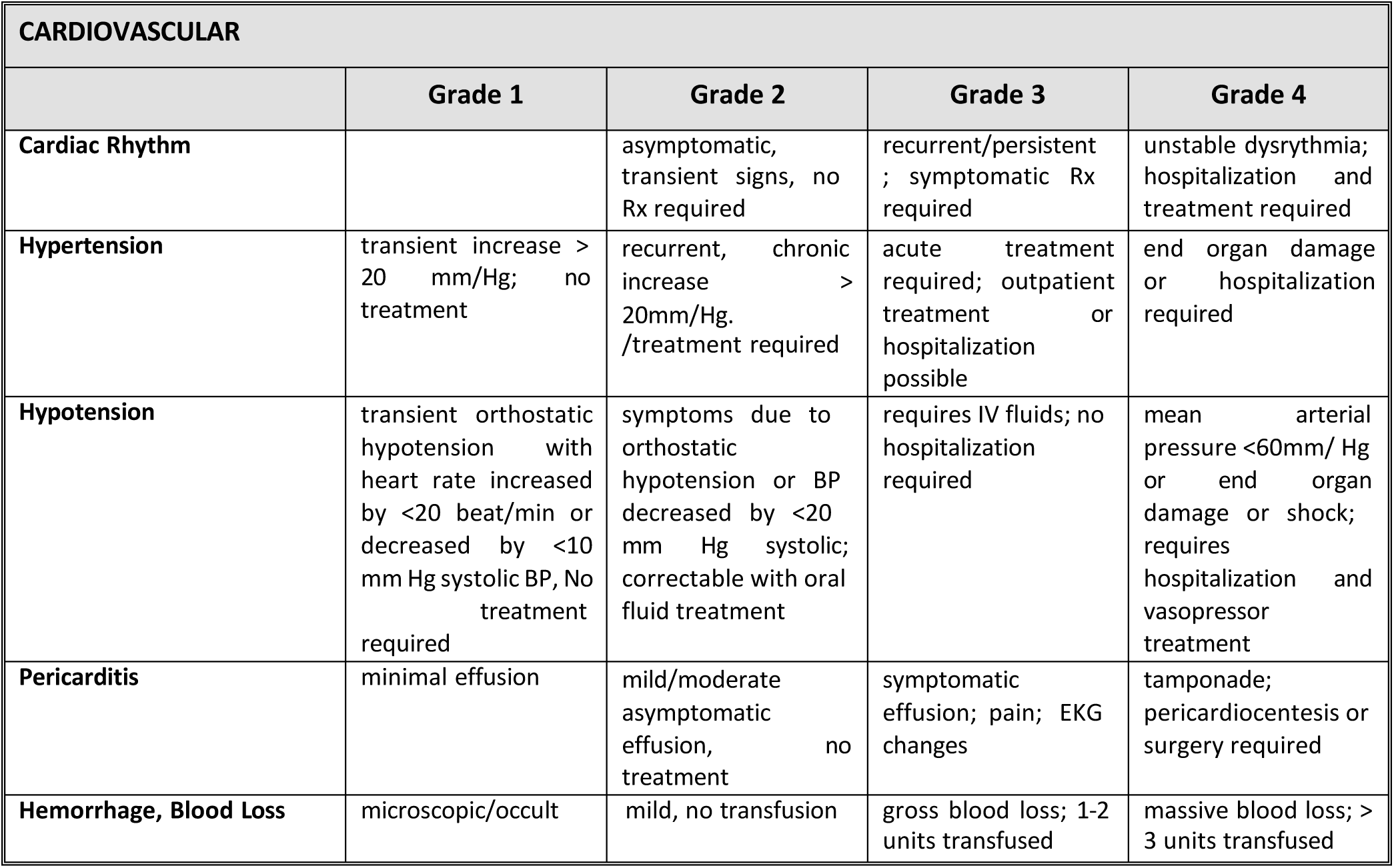
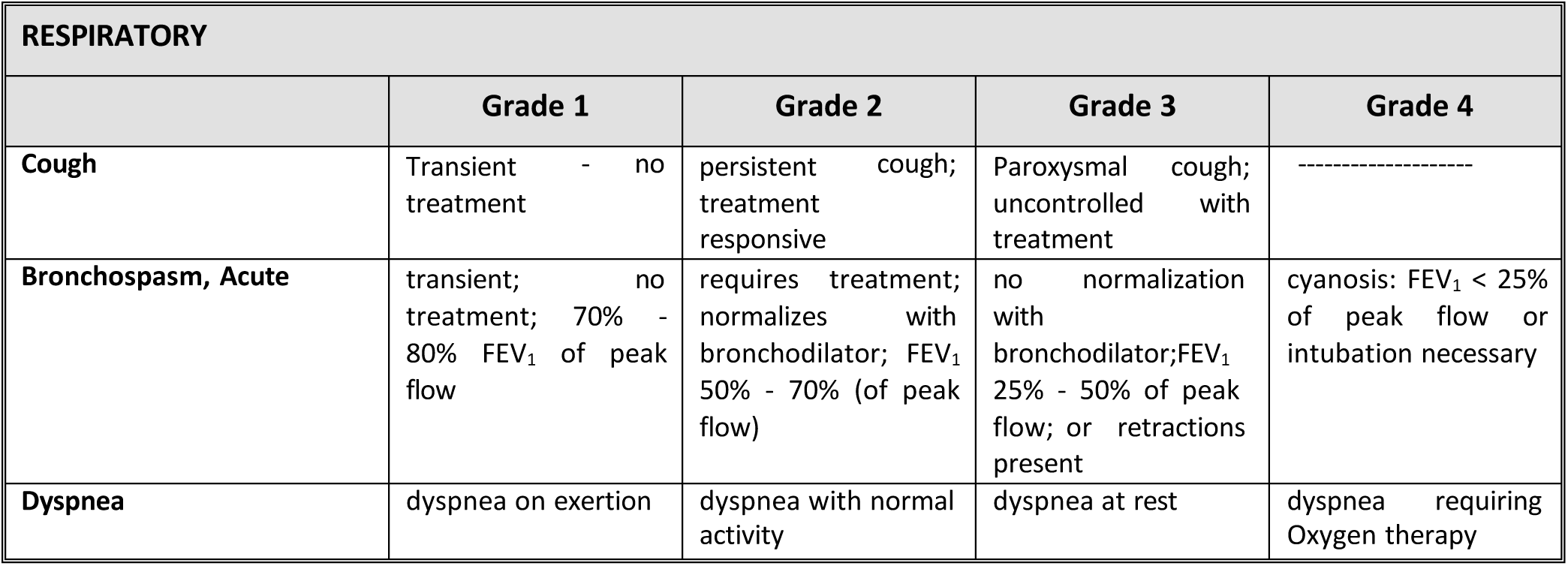
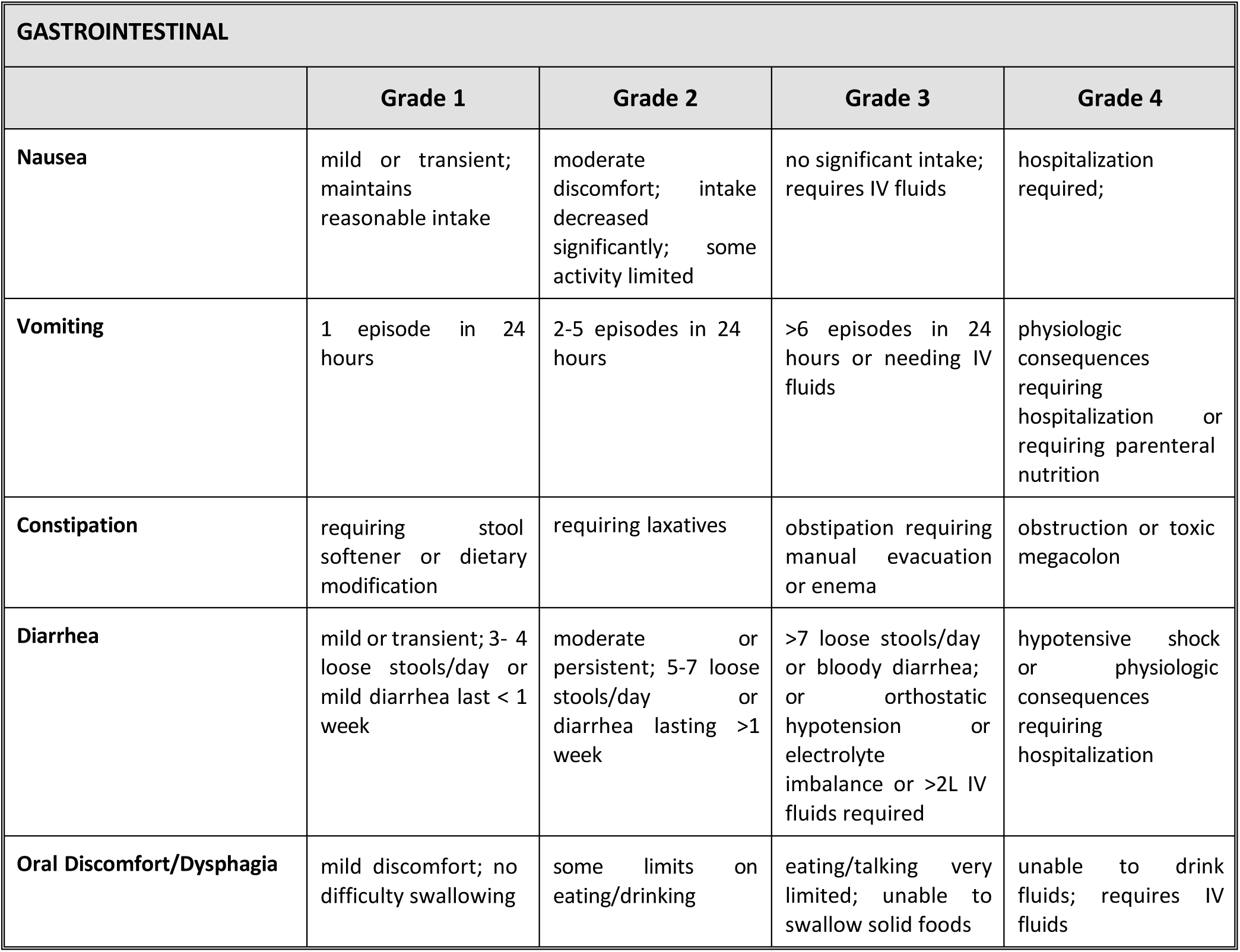
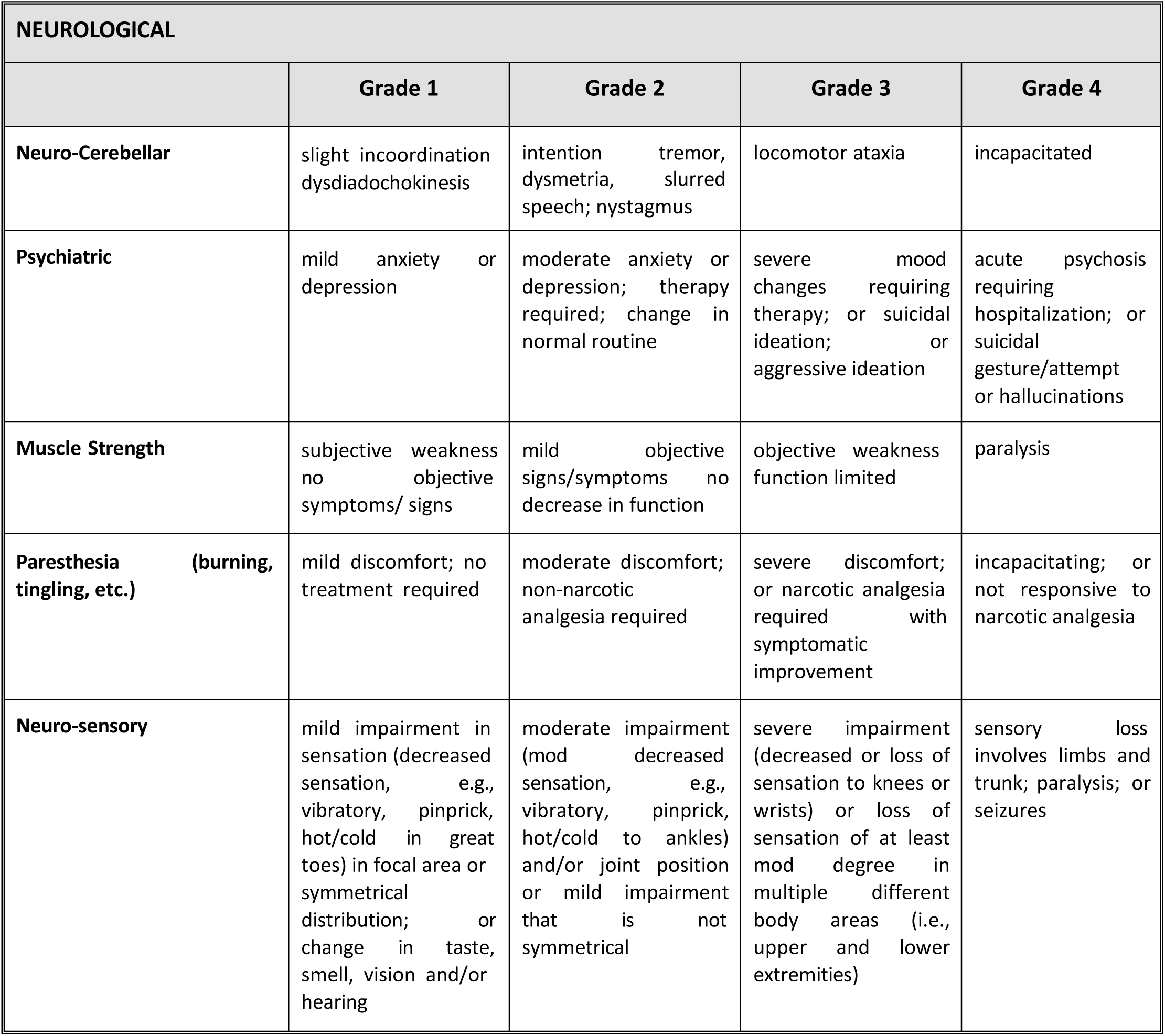
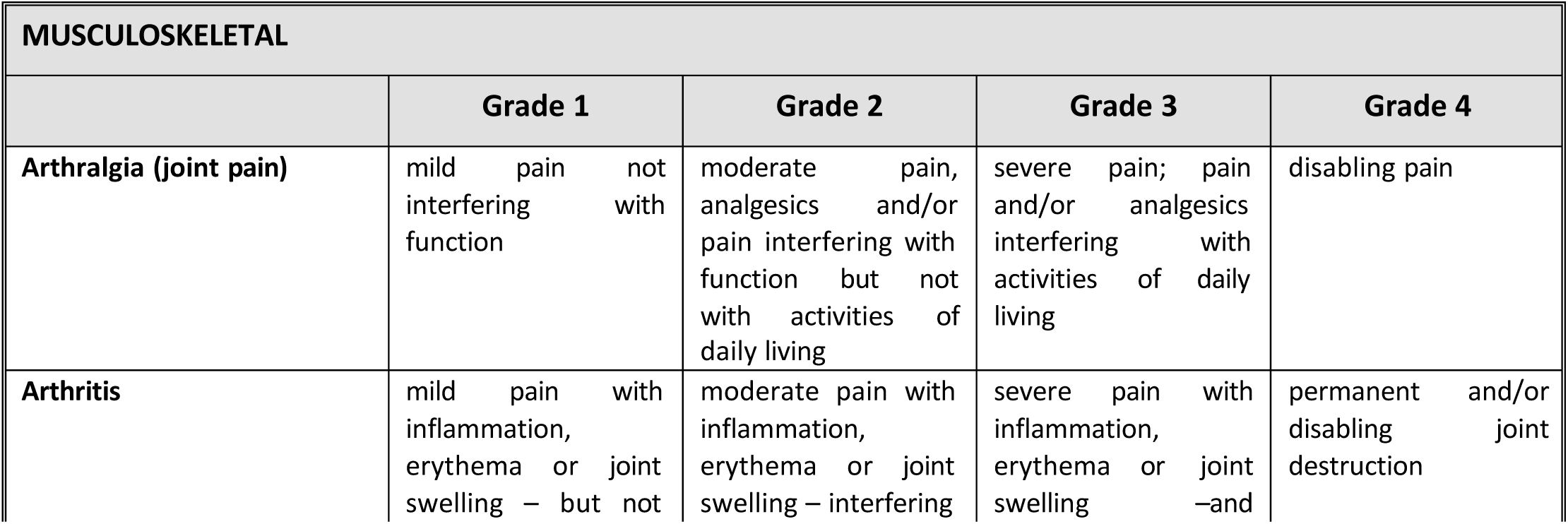
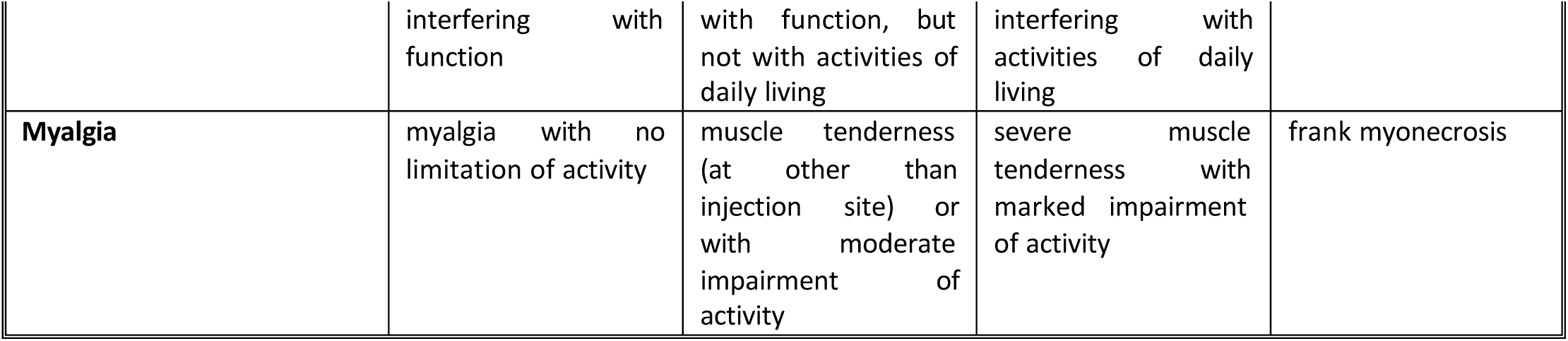
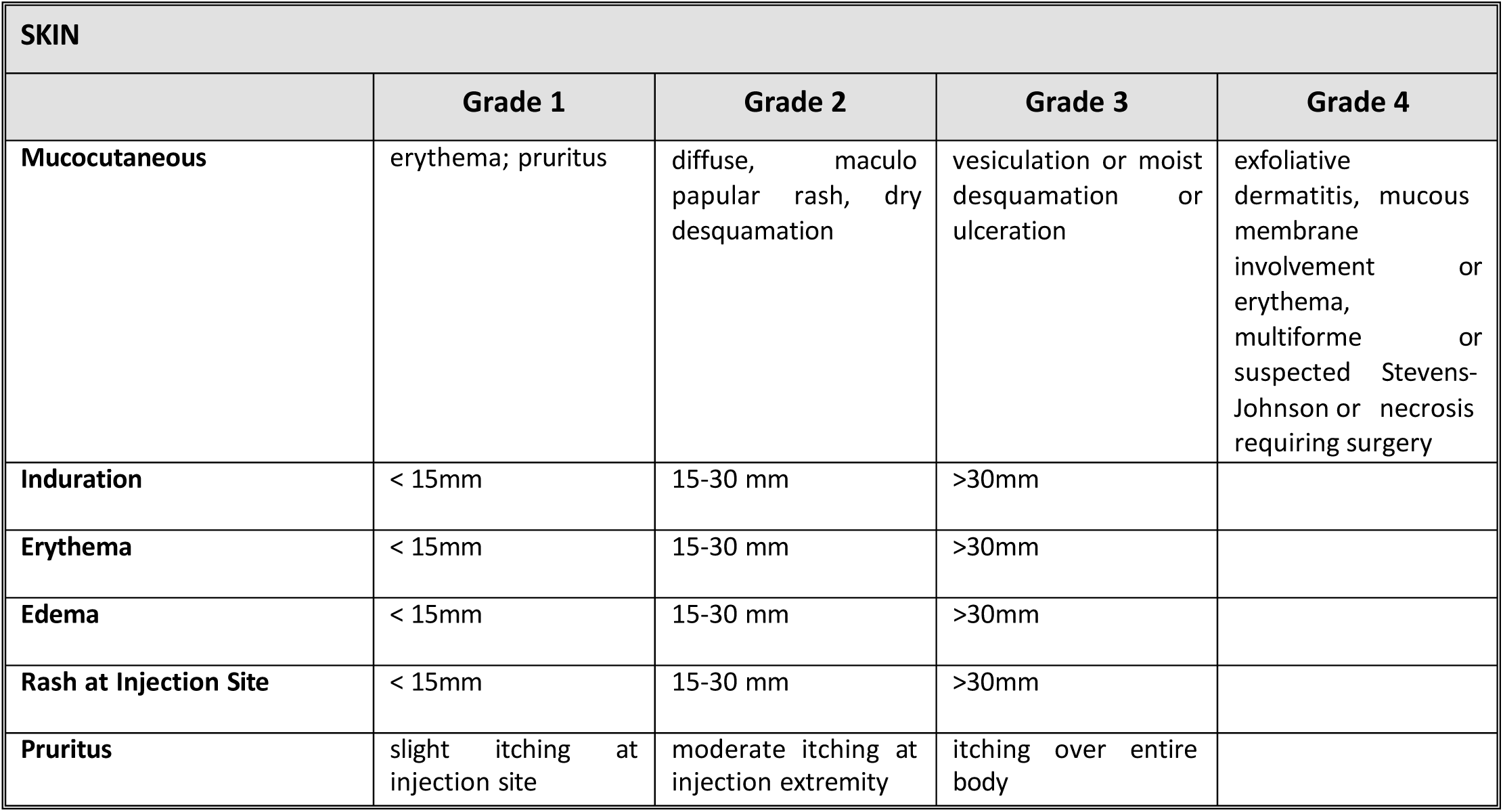
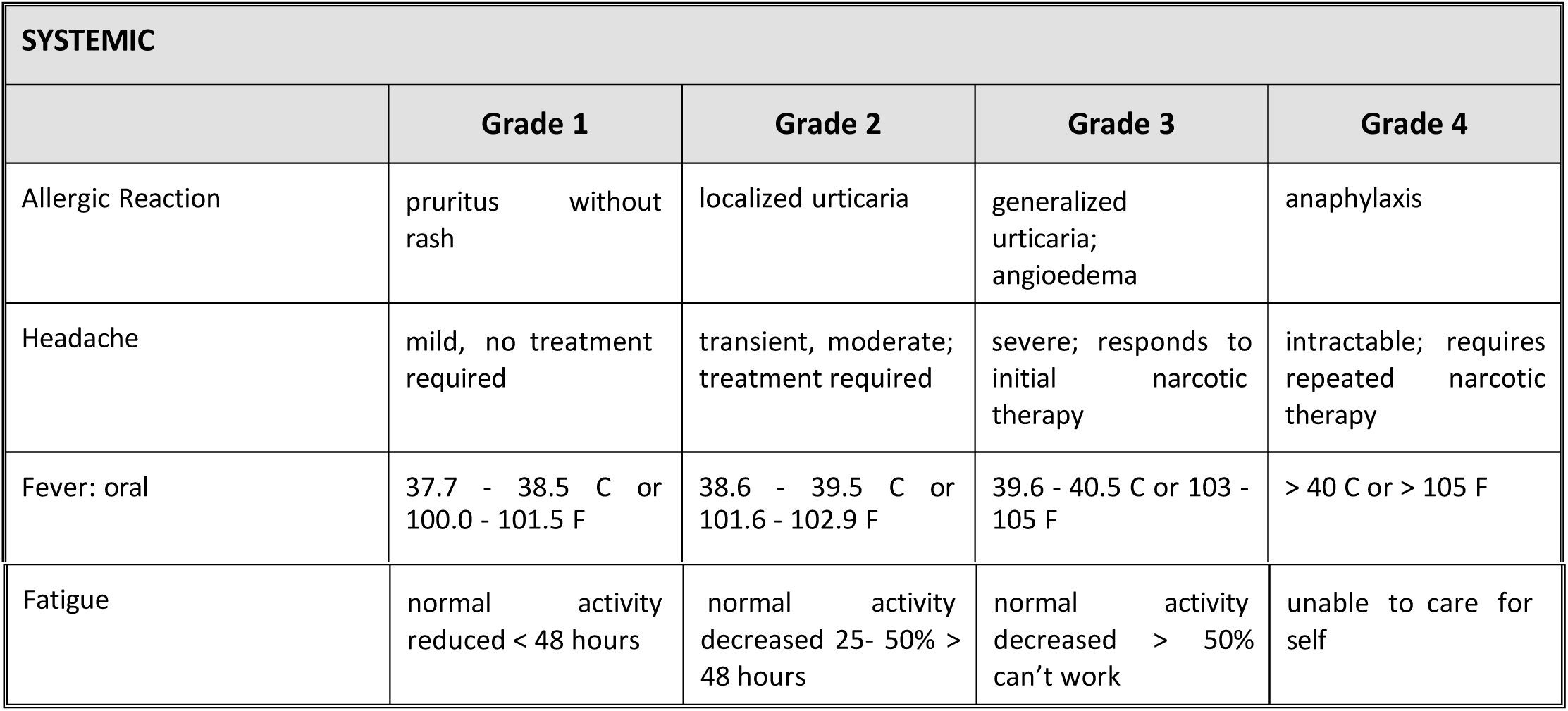

